# Computational flow cytometry immunophenotyping at diagnosis is unable to predict relapse in childhood B-cell Acute Lymphoblastic Leukemia

**DOI:** 10.1101/2024.04.18.24306015

**Authors:** Álvaro Martínez-Rubio, Salvador Chulián, Ana Niño-López, Rocío Picón-González, Juan F. Rodríguez Gutiérrez, Eva Gálvez de la Villa, Teresa Caballero Velázquez, Águeda Molinos Quintana, Ana Castillo Robleda, Manuel Ramírez Orellana, María Victoria Martínez Sánchez, Alfredo Minguela Puras, José Luis Fuster Soler, Cristina Blázquez Goñi, Víctor M. Pérez-García, María Rosa

## Abstract

B-cell Acute Lymphoblastic Leukemia is the most prevalent form of childhood cancer, with approximately 15% of patients undergoing relapse after initial treatment. Further advancements depend on novel therapies and more precise risk stratification criteria. In the context of computational flow cytometry and machine learning, this paper aims to explore the potential prognostic value of flow cytometry data at diagnosis, a relatively unexplored direction for relapse prediction in this disease. To this end, we collected a dataset of 252 patients from three hospitals and implemented a comprehensive pipeline for multicenter data integration, feature extraction, and patient classification, comparing the results with existing algorithms from the literature. The analysis revealed no significant differences in immunophenotypic patterns between relapse and non-relapse patients and suggests the need for alternative approaches to handle flow cytometry data in relapse prediction.

## 1. Introduction

B-cell progenitor Acute Lymphoblastic Leukemia (BCP-ALL) is the most prevalent pediatric cancer, impacting approximately 40,000 children globally each year. Recent clinical trials report survival rates exceeding 90% [1]. However, the remaining 15% experience relapse or refractory disease, with this subset facing a significantly worse prognosis [2]. The advancements in overall survival over the past decades can be attributed to the implementation of intensive multi-agent chemotherapy regimens tailored to specific risk groups. These groups are identified through cytomorphology, molecular biology, cytogenetics, and immunology [3]. Despite these strides, the latest data suggests that improvements in overall survival will not be reached by further adjusting regimes or incorporating novel chemotherapeutic agents. Instead, hopes for finally achieving a manageable disease lie in immunotherapies for relapsed patients and refined risk stratification criteria at diagnosis [4]. New strategies are therefore necessary to identify and select patients unresponsive to standard chemotherapy and who are at a heightened risk of relapse, given the inaccuracies of current risk allocation schemes [5]. Quantitation of minimal residual disease levels early during therapy, either by flow cytometry (FC) or by clonospecific quantitative Polymerase Chain reaction(qPCR), has been consistently reported as a major prognostic factor [6, 7]. Despite the fact that FC generates an extensive dataset of single-cell information, it is currently not utilized in risk stratification. In other words, the immunophenotype of the leukemic clone at diagnosis lacks prognostic value. Several factors impede the comprehensive exploitation of this type of data. One of them is the inherent challenge of managing high-dimensional data, especially in the clinical setting [8]. Another reason is the difficulty in gathering a sufficiently large retrospective cohort of patients. Indeed, the lack of prognostic value means that they are less frequently published than other clinical and pathologic information and therefore stored more casually. Lastly, despite ongoing efforts to standardize instruments and protocols [9, 10], differences in adherence to standards, cytometer settings, and calibration continue to pose significant challenges for multicenter data integration [11].

The recent emergence of computational flow cytometry [12] has paved the way for automated and more thorough analyses of this type of data. This interdisciplinary field brings together flow cytometry with modern pattern recognition and statistical techniques for data processing and analysis. In combination with machine learning, these techniques can be applied for survival or relapse prediction, sample classification, or subpopulation detection [13]. Surprisingly, there is a notable lack of applications of these tools in the context of BCP-ALL, with only a few published works. For instance, a study by Reiter et al. [14] gathered a dataset of 337 bone marrow samples and employed supervised machine learning to automate minimal residual disease assessment on day +15. Good et al. [15] compiled data from 54 patients and developed a classifier that organized cells based on developmental stage and achieved a high accuracy in relapse prediction [15]. Two additional preliminary works from our group complete this landscape [16, 17], one based on percentile differences of marker expression and the other on topological data analysis. There are other works focused on relapse prediction but without employing FC data [18, 19].

In this work, we set out to fill this gap and determine whether standard flow cytometry panels at the time of diagnosis contain prognostic information. To this end we collected the largest database of FC data of children with BCP-ALL for a computational analysis yet. We integrated tools from computational flow cytometry for data preprocessing and normalization and designed a comprehensive pipeline for feature extraction and classification. We identified cellular subpopulations across the cohort of patients and we assessed the prognostic value of cell abundance and marker expression with a variety of metrics. We additionally contrasted our results with other algorithms for biomarker discovery already presented in the literature. Contrary to our initial hypothesis, our results dismiss the utility of differential expression and distribution-based feature engineering for FC-based classification. We conclude the study by offering insights into the absence of discernible differences between relapse and non-relapse patients and proposing potential avenues for further exploration in this line of research.

## 2. Materials and Methods

### Study population

252 patients from three different Spanish hospitals participated in this study. We collected data from 116 patients from Hospital Niño Jesús, Madrid (HNJ), diagnosed between January 2013 and January 2022; 80 patients from Hospital Virgen de la Arrixaca, Murcia (HVA), diagnosed between May 2011 and July 2022; and 56 patients from Hospital Virgen del Rocío, Sevilla (HVR), diagnosed between January 2012 and July 2021. 207 patients had long-term remission and 44 patients relapsed. All patients are in the age range 0-19. We dropped those which continued treatment at another institution or that had not reached 1 year of follow up, with 211 patients finally proceeding to the main analysis (Figure S1). The data collected included FC files from bone marrow samples at diagnosis and additional clinical information: Age, sex, phenotype, risk group, CNS involvement, absolute lymphocyte count (ALC), immunophenotype and genetic information (karyotype and chromosomal translocations). Informed consent was obtained from the parents or legal guardians according to the Helsinki Declaration.

### Treatment

Treatment was administered according to the Spanish National protocols SEHOP-PETHEMA 2013 and INTERFANT-06 in patients under 1 year old. Older patients from HVR and HVA followed the previous consecutive versions of this protocol (LAL/SEHOP 01 for low risk patients, LAL/SEHOP 96 for intermediate risk patients and LAL/SHOP 05 for high risk patients). These protocols are based on the Berlin–Frankfurt–Munster (BFM) backbone and consists of a four-drug induction phase, followed by a second induction phase, consolidation, reinduction, and maintenance. High risk patients receive three specific high-risk blocks, three reinduction cycles, and maintenance. The total duration of therapy is 2 years.

### Risk stratification

Risk stratification criteria is based on age, lymphocyte count at diagnosis, extramedullary infiltration, cytogenetics and early response to treatment. SEHOP-PETHEMA 2013 assigns a low risk to patients who meet the following criteria: Age between 1 and 10 years, ALC less than 20 *·* 10^9^ cells/liter at diagnosis, absence of CNS or testicular infiltration, high hyperdiploidy or presence of t(12;21), absence of t(1;19), no MLL rearrangement, good early response and good response to prednisone. High risk patients verify at least one of the following: presence of t(4;11), hypodiploidy, BCR-ABL rearrangement or poor early and prednisone response. Patients who do not meet either criteria are assigned to intermediate risk [20].

### Patient outcome

Patients are assigned to either relapse or non-relapse group. Bone marrow relapse is diagnosed with the same criteria as the initial diagnosis: presence of >25% of leukemic blasts in bone marrow. Extramedullary relapses require a biopsy of the tissue or a sample of cerebrospinal fluid for confirmation. For a patient to be included in the non-relapse group we require at least one year of disease-free survival after treatment.

### Flow cytometry data

All data is retrospective. Bone marrow samples have been handled following standard clinical procedures (there is no specific design for this study). Monoclonal fluorochrome-conjugated antibody combinations employed at each hospital are shown in Table S1. Some patients presented variations from this standard (marker changes, additions or omissions). A visualization of all markers per patient is provided in Figures S2 and S3.

### Preprocessing of flow cytometry data

Preprocessing encompassed a manual and a computational step. The manual step consisted in checking each aliquot for acquisition errors and removing doublets and debris (Figure S4A). At this step we required that all aliquots contain CD19 and CD45 markers. For this reason, certain patients (mostly those diagnosed at earlier dates) were excluded from the study (1 from HVA and 7 from HVR). Aliquots with too little cells or with strong batch effects were also removed.

The compensated files were subsequently exported to undergo the computational preprocessing step [21]. This preprocessing involved transforming data with the standard logicle transform, removing margin events (this is done more efficiently here than manually) and renaming the channels to uniformize marker names across patients. Finally, each marker was normalized to the [0,1] interval by means of a modified max-min transformation: Instead of taking the maximum and minimum values, we took the 99^th^ and 1^st^ quantile respectively, making the normalization more robust to outliers. This transformation implies that we are comparing relative expression of a marker instead of the absolute expression.

Finally, we had to consider the issue of backbone markers displaying inter-aliquot differences. Some causes of this variability are staining problems, acquisition errors and other batch effects. To account for this source of heterogeneity we first sampled 10000 cells from each tube and then performed quantile normalization, a technique already used in RNA-seq data to make distributions more similar. Instead of normalizing the whole distribution we followed the approach in the cytoNorm algorithm [22]: we performed flowSOM clustering with 5 clusters and then normalized on a per cluster basis (Figure S4B).

### File merging

File merging (also file matching, panel merging or imputation) refers to the process of combining all the information from a FC experiment into a single file. The issue arises from the fact that flow cytometers can measure a limited number of colors, i.e. the expression of a limited number of protein cell markers. To obtain information for more markers, the sample is divided in several tubes or aliquots and each tube measures a different set of proteins, while maintaining a subset of them constant (backbone markers). This is enough for manual inspection of the sample but for data analysis the combined file allows for a much deeper analysis.

Several methods have already been developed for this purpose. Most of them rely on nearest neighbor imputation: Backbone markers are used to find the closest neighbors (cells with the highest surface protein similarity), and the missing information is copied from the respective neighbor. This was first published by Pedreira et. al. [23]. Later works use slightly modified versions that aim to correct artifacts and biases: cytoBackBone [24] includes the concept of acceptable and non-ambiguous nearest neighbors (data is only imputed if a cell’s closest neighbor is also the other cell’s closest neighbor) and CYTOFmerge [25] used median expression from the closest 50 neighbors instead of the single closest one. A more recent method (cyCombine) [26] follows a different methodology: It finds clusters in the space of backbone markers and then approximates the distribution of the remaining markers using kernel density estimation. The missing information is then imputed using probability draws. This is similar to other approach by Lee et. al. [27], which requires domain knowledge but demonstrated that pre-matching clustering enhances performance and reduces the risk of spurious cell populations appearing in the data. These previous steps improve quality of merging in terms of preserving the original distribution at the expense of removing cells that are too exclusive of one file and that would otherwise impute noise.

In light of these advances, the question arises as to which one is the most suitable method for conducting downstream analysis on a patient dataset. A recent comprehensive review delved into this question [28], using an array of metrics to compare the performance of the different algorithms. They concluded that there is not a clear winner and caution needs to be taken when performing downstream analysis with imputed data. A similar approach was carried out by Perdersen et. al. [26] when demonstrating the cyCombine functionality. The Earth’s Mover Distance (EMD) was employed to compare the distribution of a marker in the original tube versus the merged file. This distance, also known as Wasserstein distance, measures the minimum cost required to transform one distribution into another. In the context of flow cytometry, this cost is associated with moving cells from one marker expression state to another. Lower EMD values indicate a closer match between the original and imputed distributions, suggesting a more accurate imputation process. Its suitability for comparing marker expression distributions in the context of flow cytometry was recently demonstrated [29].

Here, we preprocessed patients from each hospital as described above and imputed the missing values according to the four methods mentioned in the main text: Direct nearest-neighbor imputation (basic), CYTOFmerge, cytoBackBone and cyCombine. We computed the EMD between the expression in the merged file and the expression in the specific aliquot in which they were present. We also obtained visualization of marker intensity in both files as well as bidimensional plots to assess the quality of the merging routine.

### Clustering and visualization

In the context of computational flow cytometry, clustering algorithms are used to detect cell populations for downstream analysis. These cell populations are then compared across patients or time points. A wide variety of algorithms of this kind are already available in the literature, and their performance has been compared through the FlowCAP challenges [30]. For our work we selected the FlowSOM algorithm, which reported the best performance and runtime in a comprehensive benchmarking study [31].

FlowSOM algorithm first clusters the data on a higher resolution (clusters) and then obtains an optimal lower number of metaclusters by aggregating with consensus clustering. Without prior knowledge about the number of groups in the data, the usual approach is to compute a metric of clustering stability in order to make an informed choice. A classical metric in consensus clustering is the proportional change in the Area Under the Curve of the cumulative distribution function [32], which measures the stability of clusters by quantifying the proportion of times pairs of items are assigned to the same cluster across multiple clustering iterations. Alternate metrics that deal with inconsistencies of the AUC have been proposed, such as the Proportion of Ambiguous Clusters (PAC) [33], which quantifies how frequently pairs of data points are assigned to the same cluster across multiple runs. In this study we used both metrics to make an argument for the optimal number of clusters. Since the FlowSOM algorithm is initialized at a random state, we performed 50 runs and compared average values. FlowSOM was run with parameters *x* = 5, *y* = 10 and *maxK* = 20. The optimal number of clusters is typically determined by a knee or elbow that can be computed automatically with the kneedle algorithm [34]. Once an optimal number of metaclusters *k_opt_*is selected, we obtained the final metacluster assignment by aggregating the results of 50 new runs of FlowSOM. This was done by minimizing the Euclidean dissimilarity across the 50 metacluster assignments.

For visualization we used the dimensionality reduction technique UMAP. This technique computes a two-dimensional representation that preserves the structure of the cell subpopulations [35]. We randomly subset 1000 cells from each patient and pool the subset files to obtain the embedding of the bone marrow of all patients. After a visual exploration, we selected UMAP hyperparameters *min*_*dist* = 0.01, *n*_*neighbors* = 15 and the rest with default values (Figure S5).

### Feature extraction

The most common features for analyzing flow and mass cytometry data are abundance (relative or absolute) and expression, measured as the median intensity of a marker (MFI), in general or on a per-cluster basis. This has been the case in most of the studies and methods used for biomarker discovery in FC data applied to leukemia (Table S2). However, a single number might not be enough to characterize the full marker distribution and thus to discover differences in expression, intensity and immunophenotype. Here, we computed for each cluster not only the abundance and median expression but also the first four moments of the distribution (mean, standard deviation, skewedness, and kurtosis). We created a dataset for each feature and a dataset with all features together, in order to find which characterization is best for detecting differences in expression and to see if the combination of all enhances the predictive capacity.

### Classification

Most of the published methods for analyzing FC data (Table S2) use linear models to perform moderated tests in order to find significant differences in expression (median intensity). The exception are neural network based algorithms, which do not explicitly perform feature selection but include the FC file as input for the algorithm. The differential expression methodology is standard in transcriptomics analysis, when looking for genes that are overexpressed under given conditions [36]. For the problem and the hypothesis of this study, finding a significantly over- or under-expressed marker might not be enough to distinguish a relapse from a non-relapse patient. In other words, while we would be able to state that relapse patients on average have a higher expression of certain marker, we would not be able to say whether a new patient belongs in the relapse or non-relapse group. Further, these analyses consider markers individually, but it could be the case that, while there might not be significant differences in MFI of a marker, we could find a region in the space of MFIs that separates both groups of patients.

Without any previous knowledge about the characteristics of this region and given that it can be quite different depending on which metric we are considering, we could not say a priori which classification model was best for this task, nor which hyperparameters of such model were optimal. For this reason, the classification routine had to include some form of internal validation to make this decision based on the data. We did this by means of nested cross-validation [37, 38]. This approach consists of two cross-validation loops, an outer loop and an inner loop. The inner loop is used to find the best model and its hyperparameters, and the outer loop is used to get an estimate of performance in unseen data. For the inner loop we performed 9-fold cross-validation repeated 20 times to get a more robust estimate, and for the outer loop we performed 5-fold cross-validation, repeated 10 times. This resampling scheme implied that each inner fold contained 16 patients on average, with 2 of them belonging to the relapse group.

We chose 4 models that are widely used and ensure that different types of boundaries are explored: K-Nearest Neighbors, Naïve Bayes Classifier, Random Forest, and Linear Support Vector Machine. Each time we trained a model we use random grid search to select the optimal hyperparameters (Table S3). The best model was selected based on the one standard deviation rule using the area under the Precision-Recall curve (AUCPR), which is more suitable for problems with unbalanced data [39]. Hyperparameter estimation and model selection were thus performed together [40].

For every dataset, the nested cross-validation routine produces 50 performance estimates (AUC-PR) and identifies 50 ‘best models’ (obtained from the 10 repetitions of 5-fold cross-validation in the outer loop). We summarized the 50 AUC-PR values by calculating their average, and the 50 best models by using a measure of heterogeneity as a surrogate of the stability of the routine. This stability measure is assigned a value of 1 if the same model is consistently selected in all outer folds, and 0 if the four models are equally frequent. It is important to note that while this measure can indicate instability or unsuccessful optimization, it is possible for two models to perform nearly equally well in identifying the best boundary, making them equally suitable for the task at hand. Thus, it is essential to consider the degree by which the top model has been selected and its associated level of performance.

To sum up, for each dataset we had a measure of the predictive information it contains (average AUCPR) and a proxy of the reliability of this measure (stability index). These two metrics were employed in conjunction to assess the predictive information across different metrics and metaclusters. Figure S6 provides an overview of the feature extraction and classification steps.

### Comparison with other algorithms

We already mentioned the existence of other algorithms and studies that aim to predict a clinical outcome from flow cytometry data (Table S2). The way they are designed follows a similar pattern: All of them begin from a set of flow cytometry files (one per patient) and cells are clustered with a different algorithm depending on the method. Each cluster is summarized by means of the abundance and the median fluorescence intensity of a marker, and these are in turn used for classification. Generalized linear models are the usual choice, as many algorithms are inspired by RNA or DNA microarray data analysis. The exception to this two-step process are neural networks based algorithms, since feature extraction is performed in the inner layers of the network. The pipeline that we followed here aimed to generalize this ‘classical’ approach by going beyond the typical characterization of a marker distribution (MFI) and by including a broader and more thorough classification routine. To validate the conclusions of this study, we selected four of the most cited algorithms and compared the results. Below we summarize the characteristics and functionality of the selected algorithms.

- Cydar [41] identifies differentially abundant cell populations between groups. It was originally proposed for mass cytometry data but can be extended to any multidimensional dataset. It clusters cells into hyperspheres, extracts cell abundance and tests for significant differences by means of a negative binomial generalized linear model, controlling for the spatial false discovery rate. In this study we subsampled 1000 cells from each patient, clustered with scaling factor 0.2, removed hyperspheres with average counts below 5 and applied the QL framework to test for significant differences. After correcting for multiple testing (spatial FDR<0.05), relevant hyperspheres and the respective fold changes in abundance were visualized on the UMAP embedding of the dataset.
- Citrus [42] identifies cell subpopulations associated with a clinical or experimental outcome. It clusters cells in a hierarchical manner, extracts either abundance or median expression and uses regularized supervised learning algorithms to identify clusters of interest. For this method we also subsampled 1000 cells from each patient. We clustered with a minimum cluster size of 5% and 5 folds and tested with the nearest shrunken centroids algorithm (PAMR).
- CellCNN [43] uses a convolutional neural network to detect rare cell subsets associated with disease. As explained above, it bypasses an explicit feature extraction process to go directly from the multicell inputs to the model prediction, drawing inspiration from multiple instance learning. We ran the convolutional neural network with 1000 cells, 1000 subsets, quantile normalization and scaling already performed and the rest of parameters with the default values. The default function performs hyperparameter tuning via a single train-test split. We further included an outer loop (20 repeats of 5-fold cross-validation) to obtain an unbiased estimate of performance, since a single train-test split would make the estimation more prone to bias.
- Diffcyt [44] employs a combination of high-resolution clustering and empirical Bayes moderated tests adapted from transcriptomics to perform differential discovery analyses. It is specifically intended for complex and/or flexible experimental designs. Like Citrus, each cluster is characterized by abundance and median marker expression and these are modelled by statistical methods based on the negative binomial distribution (Bayes estimation and generalized linear models among others). We followed a previously published workflow to run this framework [45]. We reused the FlowSOM clustering obtained in the visualization step of the study and used the edgeR method for differential abundance testing and the limma method for differential expression testing.

### Software

Manual preprocessing step was performed by means of FlowJo^TM^ v10.9 Software (BD Life Sciences). The computational step was carried out in RStudio (v2023.06.1+524, Posit team 2023) with the R Statistical Software (v4.2.2, R Core Team 2022), using packages flowCore (v2.12.2, available at Bioconductor) and flowWorkspace (v4.12.1, available at Bioconductor). File matching was also performed in R adapting the code from packages cytoBackBone (https://github.com/tchitchek-lab/CytoBackBone), cyCombine (v0.2.15, available at https://github.com/biosurf/cyCombine) and CYTOFmerge (https://github.com/tabdelaal/CyTOFmerge). Clustering and visualization made use of packages FlowSOM (v2.8.0, Bioconductor), ConsensusClusterPlus (v1.68.0, Bioconductor), clue (v0.3-65, CRAN) and uwot (v0.1.16, CRAN). Classification was performed with caret (v6.0-94, CRAN) and rsample (v1.1.1, CRAN) packages. For the other algorithms of the literature, packages Cydar (v1.24.0, Bioconductor), Citrus (v0.0.8, available at https://github.com/nolanlab/citrus) and Diffcyt (v1.20.0, Bioconductor) were run in R and cellCNN (https://github.com/eiriniar/CellCnn) was run in Python v2.7 (Python Software Foundation https://www.python.org/), all of them making use of the open source code provided at their respective websites.

### Hardware

The computational preprocessing, file merging, visualization and feature extraction routines were performed on a 3,4 GHz, 4-core, 16 GB memory iMac machine. The classification routine was run on a 3,2 GHz, 16-core, 96 GB memory Mac Pro machine. Runtime per dataset was 8-9 minutes (running each outer fold in a 31-core parallel cluster).

### Data and code availability

The source code and functions used in this article can be consulted at https://github.com/Almr95/Relapse-Prediction. This repository also includes the preprocessed and merged files of the 188 patients selected for the main analysis. The full database of anonymized FC files is available at http://flowrepository.org/id/FR-FCM-Z7A2.

## 3. Results

We collected data from 252 patients from three hospitals, diagnosed between 2011 and 2022. Risk stratification criteria, treatment protocols, and outcomes are detailed in the ‘Methods’ section. Table S4 shows their clinicopathologic characteristics. The full cohort presents a relapse rate of 17,5%, in line with recent world-wide reports [46]. Most patients present a common immunophenotype and belong to the intermediate risk group. The frequency of genetic alterations is also within common ranges reported in European countries [47]. After preprocessing, filtering and merging (see ‘Methods’ and Figure S1), 188 patients were retained for analysis. Their clinicopathologic characteristics are shown in Table 1. The only relevant differences with respect to the full cohort are a lower proportion of high-risk patients (2.7% vs. 4.0%) and a higher percentage of relapse patients (20.2% vs. 17.5%), still within reported ranges.

**Table 1:** Summary of clinicopathologic characteristics of patients retained for analysis. HVR = Virgen del Rocío Hospital, HVA = Virgen de la Arrixaca Hospital, HNJ = Niño Jesus Hopital.

|  | Dataset 1 (HVR)<br>(N=46) | Dataset 2 (HVA)<br>(N=47) | Dataset 3 (HNJ)<br>(N=95) | Total<br>(N=188) |
| --- | --- | --- | --- | --- |
| Sex - no. (%) |  |  |  |  |
| Male | 27 (58.7) | 24 (51.1) | 44 (46.3) | 95 (50.5) |
| Female | 19 (41.3) | 23 (48.9) | 51 (53.7) | 93 (49.5) |
| Age at diagnosis - yr |  |  |  |  |
| Median | 3 | 5 | 4 | 4 |
| Range | 0 - 13 | 0 - 15 | 0 - 16 | 0 - 16 |
| Long term status - no. (%) |  |  |  |  |
| Relapse | 11 (23.9) | 4 (8.5) | 23 (24.2) | 38 (20.2) |
| No relapse | 35 (76.1) | 43 (91.5) | 72 (75.8) | 150 (79.8) |
| Immunophenotype - no. (%) |  |  |  |  |
| Common | 29 (63.0) | 36 (76.6) | 88 (92.6) | 153 (81.4) |
| Pre-B | 14 (30.4) | 9 (19.1) | 4 (4.2) | 27 (14.4) |
| Pro-B | 2 (4.3) | 2 (4.3) | 3 (3.2) | 7 (3.7) |
| Mixed | 1 (2.2) | 0 (0) | 0 (0) | 1 (0.5) |
| Bone Marrow blasts<br>at diagnosis - % |  |  |  |  |
| Median | 80.4 | 78.8 | 85.0 | 81.7 |
| Range | 10.0 - 96.3 | 25.6 - 95.0 | 30.0 - 99.0 | 10.0 - 99.0 |
| Leukocytes - cell/nL |  |  |  |  |
| Median | 8.29 | 7.16 | 11.07 | 8.61 |
| Range | 1.61 - 214.21 | 0.54 - 336.19 | 0.21 - 294.0 | 0.21 - 336.19 |
| Central Nervous System<br>involvement - no. (%) |  |  |  |  |
| Yes | 2 (4.3) | 2 (4.3) | 10 (10.5) | 14 (7.4) |
| No | 44 (95.7) | 45 (95.7) | 85 (89.5) | 174 (92.6) |
| Risk at diagnosis - no. (%) |  |  |  |  |
| High | 1 (2.2) | 3 (6.4) | 1 (1.1) | 5 (2.7) |
| Intermediate | 20 (43.5) | 24 (51.1) | 76 (80.0) | 120 (63.9) |
| Low | 25 (54.3) | 20 (42.5) | 18 (18.9) | 63 (33.4) |
| Karyotype - no. (%) |  |  |  |  |
| High hyperdiploidy (>50) | 12 (26.2) | 2 (4.2) | 12 (12.6) | 26 (13.8) |
| Hyperdiploidy (47-50) | 3 (6.5) | 1 (2.1) | 10 (10.5) | 14 (7.4) |
| Normal (46) | 16 (34.8) | 7 (14.9) | 40 (42.1) | 63 (33.5) |
| Hypodiploidy (40-45) | 2 (4.3) | 0 (0) | 5 (5.3) | 7 (3.7) |
| Low hypodiploidy (<40) | 1 (2.2) | 0 (0) | 0 (0) | 1 (0.6) |
| No metaphases | 11 (24.0) | 6 (12.8) | 26 (27.4) | 43 (22.9) |
| No information | 1 (2.2) | 31 (66.0) | 2 (2.1) | 34 (18.1) |
| Chromosomal alterations - no. (%) |  |  |  |  |
| ETV6/RUNX1 t(12;21) | 7 (15.2) | 10 (21.3) | 24 (25.2) | 41 (21.8) |
| TCF3/PBX1 t(1;19) | 1 (2.2) | 1 (2.1) | 4 (4.2) | 6 (3.2) |
| MLL rearrangement | 4 (8.7) | 1 (2.1) | 1 (1.1) | 6 (3.2) |
| BCR/ABL1 t(9;22) | 0 (0) | 0 (0) | 2 (2.1) | 2 (1.1) |
| No alterations | 32 (69.6) | 34 (72.3) | 63 (66.3) | 129 (68.6) |
| No information | 2 (4.3) | 1 (2.1) | 1 (1.1) | 4 (2.1) |

### Integration of multi-center, multi-sample flow cytometry data

The cornerstone of the study is FC data at diagnosis. The joint analysis of multicenter data presents several challenges that needed to be addressed prior to the classification part of the study. Although FC panels for BCP-ALL are now standardized [10], we needed to account for differences arising from the use of different cytometers, changes in machine calibration with time and other batch effects. Furthermore, due to the maximum number of fluorochromes that can be used in a single experiment, each patient’s sample is split in different tubes or aliquots that needed to be integrated if all protein markers were to be analysed together (Figure 1A).

**Figure 1:**
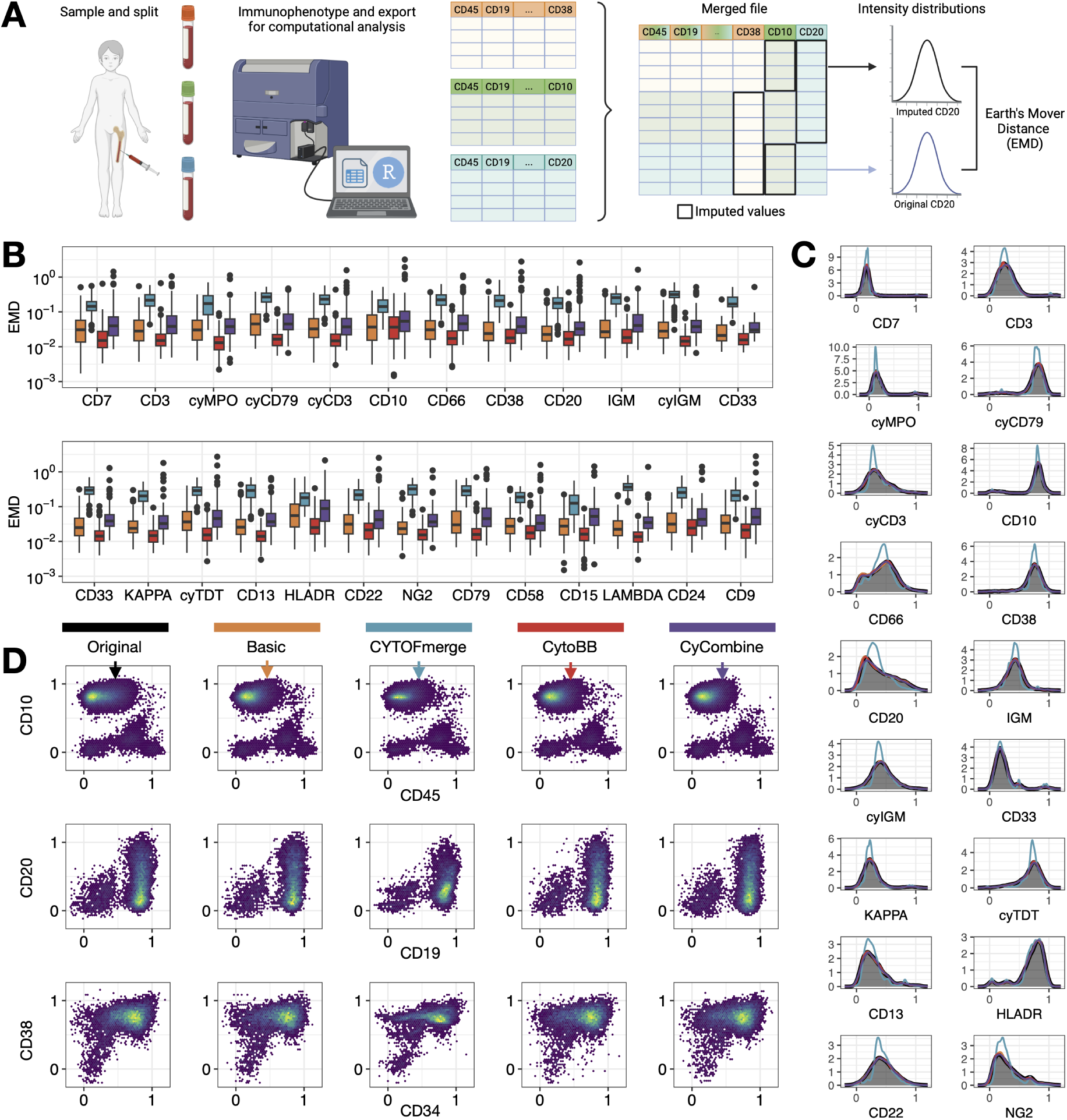
Comparison of file merging methods. **A.** Overview of the steps from data collection to merging quality assessment. Bone marrow samples are split into different aliquots and processed independently by the flow cytometer. The resulting spreadsheets are imported into RStudio and merged as described in the main text. Intensity distributions of original vs. imputed data are then compared with Earth’s Mover Distance (EMD). **B.** Boxplots summarizing the distributions of EMD for all patients and each imputed marker. We only display markers that are present in more than 50% of patients. The box includes median (horizontal line) and interquartile range (IQR). Color palette is as in [28] for comparison. **C.** Normalized intensity distributions of all markers in an example patient (HNJ_001). We show the distribution in the original file and the distribution after each imputation method. **D.** Visual inspection of the result of the imputation method for combinations of backbone markers (CD45, CD19, CD34) and imputed markers (CD10, CD20, CD38), for an example patient (HNJ_001). Plots for the remaining patients are included in Figures S7-S12.

These sources of inter-center and inter-aliquot heterogeneity were addressed here by means of a modified min-max transformation and a quantile normalization step (Figure S4, see ‘Methods’). As for the combination of several FC files into a single file, various methods have already been developed, relying mostly on nearest neighbor imputation and clustering-based imputation. In order to choose the most suitable method we used the Earth Mover Distance (EMD) to compare the distribution of a marker in the original tube versus the imputed file [29], following a recent review on the topic [28]. We compared the basic approach [23] (direct nearest neighbor imputation) with the algorithms cytoBackBone [24] (non-ambiguous nearest neighbor imputation), CYTOFmerge [25] (median of 50 nearest neighbor imputation) and cyCombine [26] (imputation by drawing from probability density estimates). Figure 1B shows the EMD of all patients for each method and each marker. For our dataset, we reached the same conclusion as in [28], with cytoBackBone consistently performing better than the other methods. When inspecting individual patients, we saw that normally cytoBackBone, cyCombine and the basic approach preserved the intensity distributions, with higher deviation for CYTOFmerge. Occasionally, cyCombine deviated as well, while cytoBackBone and the basic approach were always close. Figure 1C shows this for an example patient (HNJ_001). The rest are provided as Supplementary Material. Finally, it was also important to visualize backbone markers together with imputed markers in order to identify potential artifacts or deviations that are not visible in distributions alone. This is shown in Figure 1D for an example patient. The remaining patients are shown in Figures S7-S12. We again observed similar features to the ones described in [28]; the basic approach was prone to repeatedly imputing the same values and CYTOFmerge tended to compress or shrink the distributions, while cyCombine and cytoBakcBone yielded a more similar representation. In the light of these results, we chose to continue the analysis with the cytoBackBone method and repeat it with the cyCombine method in order to confirm the stability of the results and test the influence of the preprocessing routine.

### Cell subpopulation structure across patients

After preprocessing and file merging, we needed to select a common set of markers for study. Due to differences in extraction date, number of cells, and institution, not all markers were present in all patients. This can be seen in Figures S2, S3 and Figure 2, where we visualize the tradeoff between number of markers and number of patients. In the light of this we generated two groups, one with a higher number of patients but less markers (Selection A) and another with less patients but a higher number of markers (Selection B). The set of FC markers in group A included B-cell markers CD19, CD10 and CD20; pan-leukocyte markers CD45 and CD38; hematopoietic stem cell marker CD34 and myeloid markers CD58 and CD66c. Group B additionally included B-cell markers IGM and CD22; T-cell markers CD3 and cyCD3; myeloid markers CD13 and MPO and progenitor marker TDT, at the cost of 30 patients.

**Figure 2:**
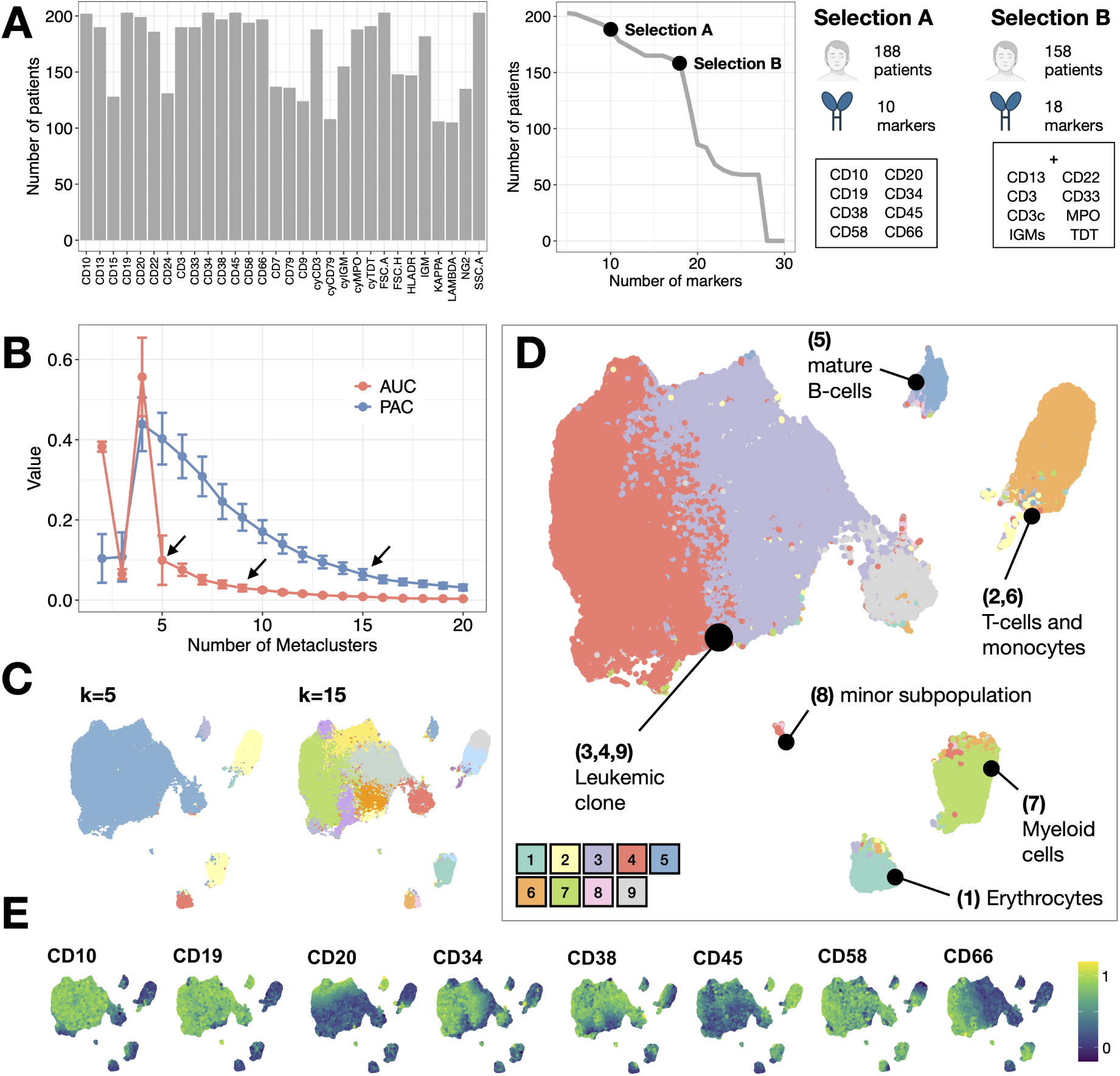
Clustering and visualization of flow cytometry data. **A.** Number of patients each marker is present in. We first show each marker individually and then the variation of total number of patients as the marker count increase. This inverse relationship or tradeoff motivates the selection of two groups of patients for study. **B.** Values of AUC (Area Under the Curve of the empirical distribution function of a consensus matrix) and PAC (Proportion of Ambiguous Clustering) according to number of metaclusters selected by FlowSOM algorithm in Selection A. **C.** UMAP visualization of 5 metaclusters and 15 metaclusters in Selection A. **D.** UMAP visualization of 9 FlowSOM metaclusters in Selection A, manually labelled according to marker expression. **E.** Relative intensity of marker expression in the UMAP embedding employed for visualization.

The next step was to visualize the structure of the bone marrow of all patients. Cell subpopulations can be obtained by means of clustering techniques, which replace the traditional manual analysis or ‘gating’ [31]. Here, we pooled all the files together and clustered via FlowSOM [48]. This algorithm obtains an optimal number of metaclusters *k* by aggregating with consensus clustering, although this optimal number can be affected by the stochasticity in the implementation of the algorithm. For this reason we performed 100 runs of FlowSOM and compared metacluster optimality through proportional change in Area Under the Curve of the empirical distribution function of the consensus matrix (AUC) and with the Proportion of Ambiguous Clusters (PAC) (See ‘Methods’). This is shown in Figure 2B for group A of patients. There is a clear elbow in AUC at *k* = 5 but not in PAC, which becomes more stable from *k* = 15 onwards. We chose to compare these two values, as well as an intermediate value of *k* = 9 that was selected automatically with the kneedle algorithm [34]. The final metaclustering assignment was selected by aggregating the results of 50 new iterations of the FlowSOM algorithm, providing more robustness to the output (Figure S13).

To visualize the clustering information on a single-cell level we obtained a 2-dimensional representation of the FC data through UMAP. The results for the reduced set of markers (Selection A) are shown in Figure 2C-D. Each cell is colored according to FlowSOM metacluster. We judged that for *k* = 5 and *k* = 15 the data was overclustered and underclustered respectively (Figure 2C), while *k* = 9 captured better the overall structure of the data as displayed in the low-dimensional UMAP space (Figure 2D). Relative intensity of marker expression is included in Figure 2E, allowing us to manually annotate the clusters found by the robust FlowSOM clustering. There were three metaclusters (3 and 4) that comprised most of the CD19+ cells and that we identified with the leukemic clone. These are immature B-cells with intermediate expression of CD45 and heterogeneous expression of CD34 and CD38. The two metaclusters were distinguished by relative expression of CD66c. We also assigned metacluster 9 to the leukemic cell population, distinguished from the other two by a negative expression of CD10. These metaclusters contained the majority of cells since the bone marrow of BCP-ALL patients at diagnosis is almost fully invaded. Metacluster 5, with a high expression of CD45 and CD20, represents healthy, mature B-cells. The remaining metaclusters represent other bone marrow cell types, including T-cells and monocytes with high expression of CD45 (metaclusters 2 and 6), myeloid subpopulations (metaclusters 7 and 8) and erythrocytes. While these subpopulations are less tracked in B-cell malignancies, here we also explored them for prognostic value. Figure S14 contains the usual two-dimensional representation of these markers for manual annotation.

Finally, for selection B (more markers and less patients) we clustered with the same markers as selection A, and the results are shown in Figure S15. The optimal number of metaclusters is 6 in this case, and we also compared with 15 metaclusters and with the optimal between both. Cell subpopulations were annotated as described above.

### Relapse prediction through cells per metacluster

In the light of the clustering result, the first question we asked was whether any of the cell subpopulations found were more associated with relapse patients. To check this we first extracted the number of patients that are included in each of the metaclusters, for the three clustering results displayed in Figure 2 (Figure 3A). Most metaclusters included all patients in the dataset, maintaining the baseline proportion of relapse patients (20.2%). Only minor subpopulations were lacking in some patients, especially in the higher resolution clustering (*k* = 15), without any particular cluster being dominated by either relapse or non-relapse patients. To investigate the predictive power of the bone marrow composition, however, we had to check not only the number of patients per metacluster but also how many cells each patient contributed with. The idea was to test if relapse patients tended to participate more in a subset of clusters, or if instead all patients contributed equally. To do so, we calculated the percentage of cells per cluster for every patient, and the results are shown in Figure 3B, again for the three possible clustering results previously discussed. No metacluster exhibited statistically significant differences (p<0.05, two-sided Kolmogorov-Smirnov test).

**Figure 3:**
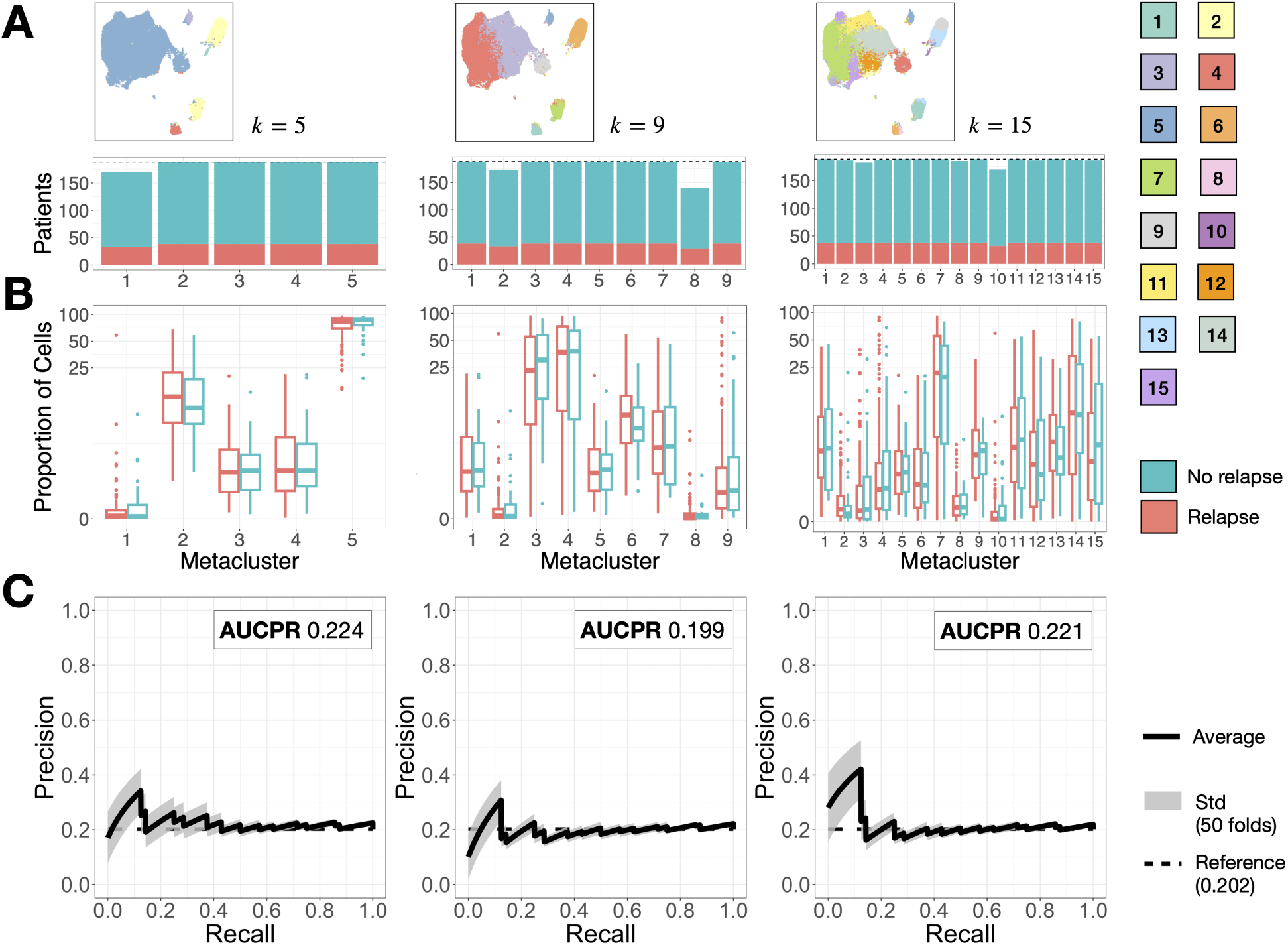
Results of abundance-based classification. **A.** Number of patients included in each metacluster, for the three selections of optimal number of metaclusters (Figure 2).**B.** Comparison of cell percentage per cluster between relapse (R) and non-relapse (NR) patients. Boxplot includes median and IQR. The scale has been transformed with an inverse hyperbolic sine for clarity. **B.** Classification results in terms of Area Under the Precision Recall Curve using information from all metaclusters. The shaded region represents the standard deviation of 50 repetitions of the classification routine (10 folds + 5 repeats). Horizontal dashed line represents the baseline precision, which equals the proportion of relapse patients in our dataset (Table 1).

This, however, was insufficient to conclude the lack of predictive power of the number of cells per cluster. Indeed, although each metacluster individually did not present clear differences, non-linear interactions between all metaclusters could create a region in which relapse patients are more clearly distinguished. We tested this by building a classifier for relapse prediction that used cells per cluster as input. We implemented a nested cross-validation scheme and included four supervised machine learning algorithms: Naive Bayes, Random Forest, K-Nearest-Neighbors and linear Support Vector Machine. For robustness, we repeated the classification 10 times. More details about the classification routine can be consulted in the ‘Methods’ section. The average Precision-Recall curves obtained for the three cases in discussion are shown in Figure 3C. We used the Area Under the Precision-Recall Curve (AUCPR) to summarize the result. This is equivalent to the average precision of the classifier and can be interpreted as the probability that a predicted relapse is a true relapse. Its value was close to the baseline precision, which is the proportion of relapse patients in our dataset (0.202, Table 1). This means that the features used for classification had no prognostic value. We finally assessed the reliability of these results by performing stability and overfitting checks (Figure S16).

To further establish this conclusion, we repeated the same analysis for group B of patients, which included more markers (IgM, cyTDT, cyMPO, cyCD3, CD13, CD22, CD3, CD33) although less patients (from 188 to 158). The results for the three clustering results are shown in Figure S17, with similar conclusions to selection A except for a slight increase in precision when using 12 metaclusters.

### Relapse prediction through relative marker expression

Following the assessment of the prognostic significance of cell abundance, we turned to marker expression within each cell subpopulation. One way to analyze this information would be to compare the intensity of the complete relapse population vs. the non-relapse population, to visualize general population-level trends. The existence of these differences, however, does not mean that each individual patient adheres to the pattern. This is shown in Figure 4A for an example metacluster in group A of patients. The distributions for the remaining metaclusters are shown in Figures S18 and S19. Most markers across the majority of metaclusters did not exhibit noteworthy disparities between the relapse and non-relapse groups. Metacluster 1 (erythrocytes) showed more expression of CD20 and CD66c in relapse patients; metacluster 2 (monocytes) displayed differences in CD34, CD58 and CD66c (this last one also in metacluster 6); and metacluster 8 (minor subpopulation) displayed lower CD20 in relapse patients. It remained to be seen that the significance of these disparities were reproducible at an individual-patient level, rather than being confined to the population level. Following the rationale of the previous section, we aimed to test whether individual patients’ marker expression could predict relapse.

**Figure 4:**
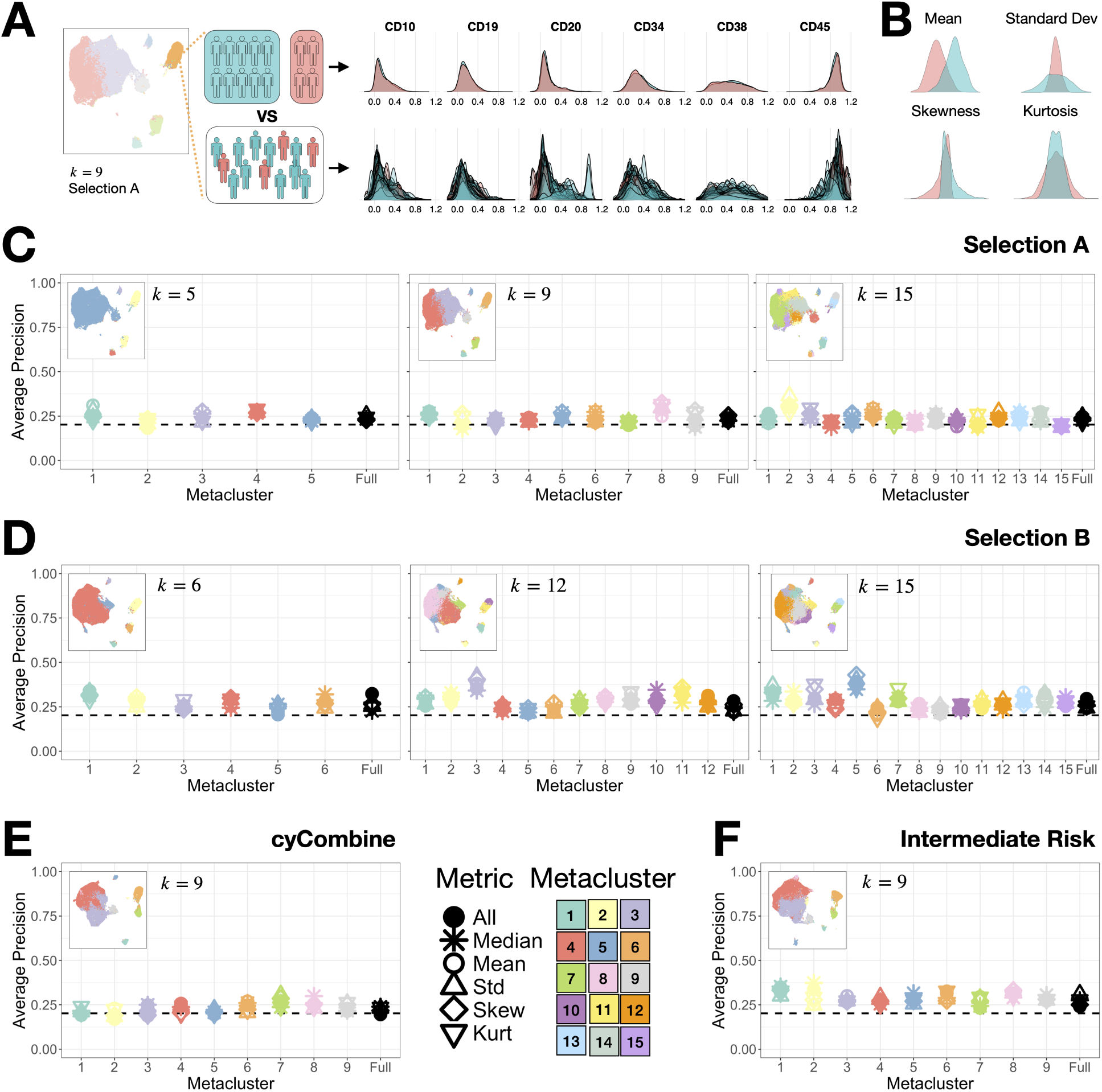
Expression-based classification. **A.** Aggregated and individual marker expression of relapse and non-relapse patients for metacluster 6 in group A of patients. **B.** Distribution metrics employed for characterizing individual patient data. **C.** Classification results in terms of AUCPR for the three clustering results in selection A. Black dashed line represents baseline precision. Color denotes metacluster. Shape denotes distribution metric. **D.** Classification results in terms of AUCPR for the three clustering results in selection B. **E.** Classification results in terms of AUCPR for a cyCombine merging algorithm (selection A, 9 metaclusters). **F.** Classification results in terms of AUCPR for intermediate risk patients only (9 metaclusters).

To address this, we first needed to summarize the intensity of expression of a marker into a single number. The usual way to do this in FC is to use the median (Median Fluorescence Intensity, MFI). Since differences in distributions can go beyond central tendency (Figure 4B), here we also considered the first four statistical moments: Mean, standard deviation, skewness and kurtosis. This procedure produced five distinct datasets, each corresponding to a specific metric. Additionally, we constructed a combined dataset with all features to explore whether a combination of metrics would yield more informative results. We used the classification routine previously described to check the predictive power of each dataset. A summary of the workflow followed in this section is shown in Figure S6. The results for the three clusterings in group A of patients are shown in Figure 4C. For each metacluster, we show the average precision (equivalent to AUCPR) obtained by each metric and metacluster. We also show the same information for the full cohort, without segregating by metaclusters. The conclusion is straightforward: the information contained in marker expression distribution lacks predictive capacity, given that the majority of AUCPRs marginally exceeded the baseline precision. We repeated the analysis with group B of patients, and the results for the three clusterings are shown in Figure 4D. The improvement with respect to the previous case was negligible. Reliability assessment for these results are included in Figure S20.

To conclude, we performed three additional analyses. We first considered whether the preprocessing of the data could be responsible for the lack of predictive information. To explore this, we replicated the analysis using the cyCombine algorithm for file matching (see ‘Methods’), and our findings concurred with the conclusions detailed earlier (Figure 4E). We then considered only those patients which were initially diagnosed as intermediate risk, to check if the more intensive treatment received by high risk patients could bias the results. This resulted in a reduced cohort of 119 patients. The results were also similar to the above (Figure 4F). Lastly, we considered wether increasing the number of cells per patient, from 10000 to 40000, would include more relevant biological information. We repeated the clustering, abundance and expression analysis, with just a marginal increase of precision when considering cell proportion per metacluster (Figure S21). Hence, irrespective of treatment received, merging technique employed, number of markers considered, number of cells per patient, cluster size and distribution metric, marker expression of FC data at diagnosis failed to predict relapse.

### Comparison with other biomarker discovery algorithms

We contrasted our findings with other algorithms from the literature designed for biomarker discovery and outcome prediction. A description of their functionality and implementation can be found in the ‘Methods’ section. The results for each of them are shown in Figure 5. The first example is Cydar [41], which is designed for differential abundance discovery. The clusters (hyperspheres in Cydar terminology) with a sufficient number of cells are projected onto the UMAP embedding employed in the previous sections (Figure 2). Those hyperspheres with significant differences in abundance (according to a lasso-regularized logistic regression) are plotted with wider radius and colored according to the fold change in abundance between both group of patients (Figure 5A). To check the predictive power of such hyperspheres, we extracted the number of cells per patient and hypersphere and ran the classification routine previously described, with results similar to the best models in the previous section (Figure 5B). The difference here is that due to the lower size of the clusters (hyperspheres), there are less patients per cluster (Figure 5C), which makes the results less generalizable. The second example is Citrus [42]. The results for both abundance and median expression (Figure 5D) indicate the lack of predictive information, regardless of regularization threshold. In both cases the null classifier (no features, largest regularization threshold) was the best classifier, with an error of 20.2%. This number is the proportion of relapse patients in our dataset, which means the algorithm was classifying all patients as non-relapse. Further, the False Discovery Rate shows all the characteristics of a classifier unable to discriminate [49]. The third example is cellCNN [43], which uses a convolutional neural network. We complemented it with a nested loop that allowed us to conclude two things: First, the lack of a inner validation routine makes the algorithm more prone to overfitting, as we see in the comparison between the accuracies of the inner and outer loops (Figure 5E). Second, the performance in terms of AUCPR did not improve previous tests (Figure 5F). Finally, we tested Diffcyt [44] on the metaclusters that were already obtained by FlowSOM (Figure 2). This algorithm showed significant differences in cell abundance in metacluster 2 (Figure 5G) and significant differences in expression in several metaclusters and markers (Figure 5H). On closer inspection, we noticed that those significant features were the ones that displayed differences in aggregated marker expression (Figure 4A and Figures S18, S19). We already discussed how this sum of distributions does not necessarily entail that individual patients follow the same trend and that a classifier could still be unable to properly predict relapse, as shown in Figure 4C.

**Figure 5:**
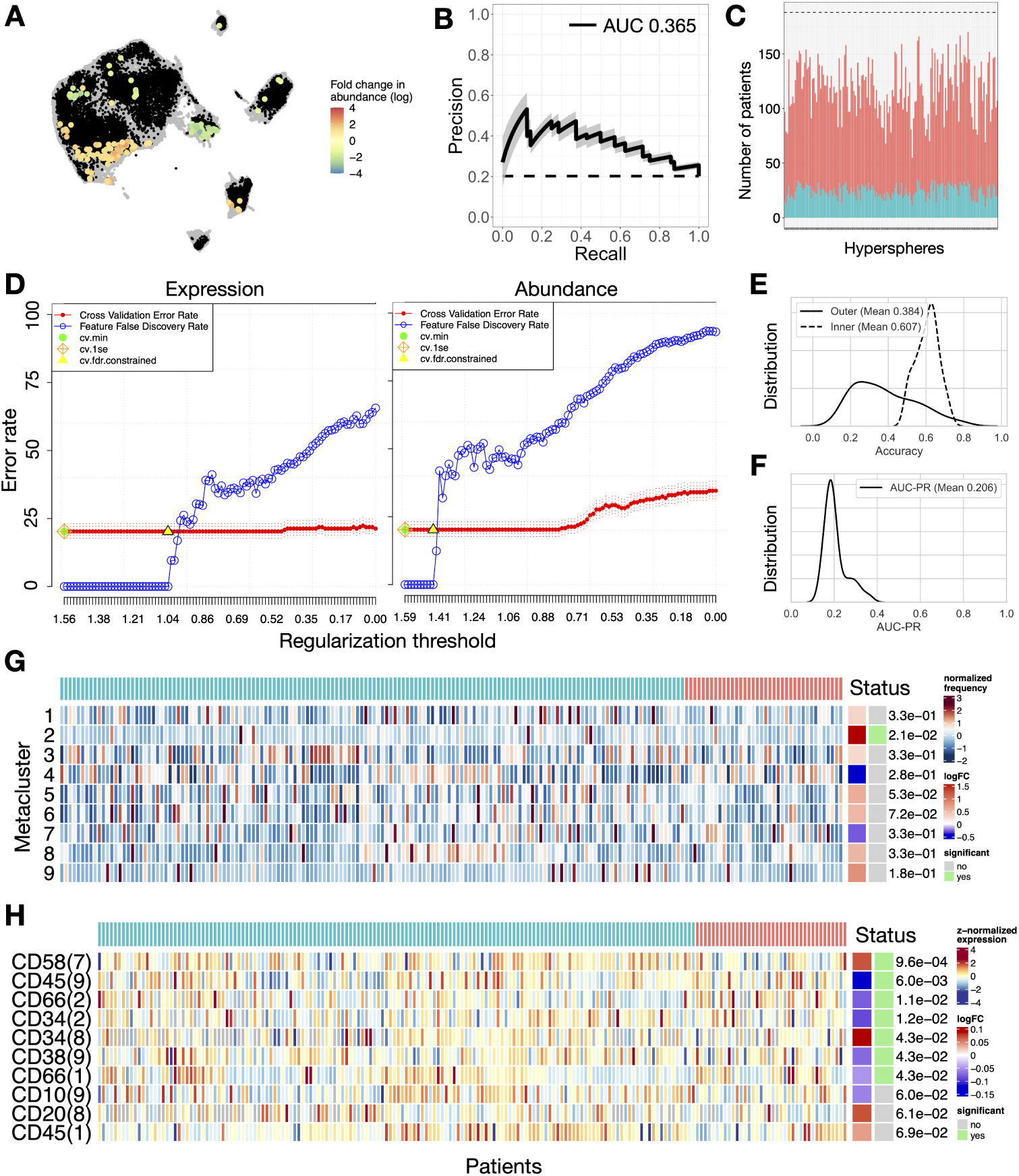
Results from other biomarker discovery algorithms. **A.** Cydar hyperspheres (black) projected on UMAP embedding from Figure 2D (gray). Significant hyperspheres are colored according to fold change in abundance. **B.** Classification results from the cell abundance of the significant hyperspheres. Interpretation is as in Figure 3B-E. **C.** Number of patients in significant cydar hyperspheres split in relapse (R) and non-relapse (NR). Dashed line displays the reference of 188 patients (38 relapses). **D.** Citrus results for median expression (left) and abundance (right). Represented are cross-validation error rate (Red) and false discovery rate (blue). Green dot represents the error rate of the best model according to the minimum cross-validation error rate. Orange rhomboid represents the error rate of the best model according to the one standard deviation criterion. Yellow triangle represents the best model according to the lowest compatible false discovery rate. **E.** Comparison between the accuracy in the outer and inner loops of the CellCNN algorithm. **F.** AUCPR curve in the outer loop of the CellCNN algorithm. **G.** Diffcyt differential abundance test. Each row contains the individual patient cell percentage in a metacluster (1 to 8). The algorithm includes the fold change between status (relapse R in red vs. non-relapse NR in blue) and the statistical significance of the results (gray vs. green) **H.** Diffcyt differential expression test. Row annotation includes the marker and the metacluster in which significant differences were found.

The conclusion of this section is that other algorithms that aim for the same goal as this study and follow a comparable methodology were also unable to detect differences between relapse and non-relapse patients. This applies to analyses centered on both cell abundance and marker expression.

## 4. Discussion

Approximately 15% of children diagnosed with BCP-ALL relapse or develop refractory disease, significantly worsening their prognosis. While advances in chemotherapy have improved outcomes, further therapeutic progress may rely on more precise risk assessments at diagnosis. In this study, we analyzed data from 188 childhood BCP-ALL patients to assess whether FC data at diagnosis could predict relapse. After preprocessing, normalizing, and integrating data, we applied FlowSOM clustering to obtain a common cell subpopulation structure across patients. We then extracted cell proportions per cluster and cluster-level features and evaluated their prognostic value using four machine learning algorithms in a nested cross-validation scheme.

The outcome of the primary analysis directly contradicts the initial hypothesis: FC data obtained at diagnosis does not appear to contain relevant information for relapse prediction. Cell abundance per cluster is unable to predict relapse, even when increasing the number of clusters. Likewise, no distribution metric is able to significantly improve the baseline precision. Considering all metrics together in a single dataset also failed to improve outcomes. Increasing the number of clusters and the number of markers, the latter with a reduction in the number of patients from 188 to 158, also yielded similar results. We repeated the analysis with a different file merging algorithm to check if preprocessing might mask differences in abundance or expression. We also restricted the analysis to intermediate-risk patients to account for the potential confounding effect of more intensive treatment in high-risk patients. Finally, we repeated the analysis with an increased cell count per patient from 10000 to 40000. The conclusion remained consistent across all these variations. The most precise classifier, achieved by increasing the number of markers and clusters, yielded an average precision of 0.37, meaning a predicted relapse has a 37% chance of corresponding to an actual relapse.

Our pipeline was designed to enhance previously published algorithms, providing a more comprehensive characterization of marker expression distributions and using non-linear classifiers with a more rigorous resampling scheme. Despite these enhancements, we verified our results against other open-source algorithms, specifically Cydar [41], Citrus [42], Diffcyt [44], and CellCNN [43]. These algorithms are widely referenced for discovery analysis in FC. Cydar identified several clusters with significant abundance differences, similar in performance to our primary analysis classifiers, though limited to a subset of the cohort. Citrus identified multiple features, but the classifier’s performance was weaker than the null model. Diffcyt identified expression differences that were significant at a population level but failed to distinguish individual patients reliably. CellCNN outcomes mirrored other classifiers, with performance marginally surpassing the baseline classifier. These findings further support our conclusion that current metrics for characterizing surface marker distributions do not differentiate relapse and non-relapse patients.

The initial hypothesis of this study rested on the premise that the leukemic clone in relapsing patients differs from that of successfully treated individuals, and that such distinctions manifest in the immunophenotype and could then be captured through FC measurements. The negative outcome we have obtained in this study offers room for diverse interpretations. It is possible that the immunophenotype of relapsing patients does not exhibit distinctive characteristics. While genetic differences are known to play a fundamental role in the origin and potentially the relapse of leukemia [50, 51], these differences may not necessarily translate to variations in surface marker expression. Rather, they may only be found through genomics, transcriptomics or metabolomics. In this line, recent research has demonstrated the feasibility of predicting relapse in infants with MLL-rearranged ALL by single-cell transcriptomics [52]. Further research is required to assess the predictive potential of a comprehensive panel of mutations for the broader population. Alternatively, distinctive immunophenotypic characteristics might emerge post-therapy. Such a scenario could be attributed to chemotherapy-induced bottleneck selection, which has been shown to impact the phenotype more significantly than genotype [53]. This could be probed by revisiting this study with FC data from a later time point, although this approach would deviate from the initial goal of refining risk stratification at diagnosis.

Another possibility is that immunophenotypic differences exist but were undetectable under this study’s conditions. Such differences may only appear in small cell subpopulations which might elude detection even with high-resolution clustering if the number of cells per patient is not increased. This hypothesis could be explored by imposing stricter limitations on the number of cells per patient, although this would inevitably reduce the total number of patients in the study. Furthermore, the file merging routines examined here have been shown to include biases in downstream analysis when the number of clusters is higher [28]. Another potential consideration is that immunophenotypic disparities manifest in markers beyond the ones routinely assessed in clinical practice. Testing this would require prospective studies and expanded antibody panels. Finally, it can be the case that immunophenotypic disparities exist but are obscured by the extensive preprocessing and normalization required to integrate data from multiple centers. No immediate alternative exists until the clinical adoption of next-generation cytometers that can measure a larger number of markers simultaneously and are more amenable to standardization. Although they were not available for our study, manually annotated populations could help in data integration, at the cost of increasing bias and reducing reproducibility [54].

Despite the scope and scale of this study, as well as the evidence gathered in support of the negative conclusion, there are still alternative ways of exploring the prognostic value of FC. We mentioned in the introduction a number of works that use FC data in the context of BCP-ALL. The closest one to the objectives of the present study is Good et al. [15], which employed mass cytometry data at diagnosis and achieved a relapse prediction AUC of 0.85 using an elastic net model. Their panels included both phenotypical and functional proteins, which supports the previous hypothesis that differences between relapse and non-relapse patients may require markers that are not used in clinical practice. As a limitation, their database only encompassed 54 patients, and the validation was confined to a single train-validation split, thereby hampering direct comparability with our results. Similar constraints apply to an earlier work by our own group that included 56 patients to identify differences in expression [16]. Given the limited success of conventional feature engineering, we recently explored the feasibility of using topological data analysis for feature extraction, obtaining high accuracy and AUC with an increased number of patients (*N* = 96) [17]. This encourages the search for differences in immunophenotype of relapsing patients by means of more complex methods.

An alternative to the above is to skip the feature extraction process altogether and allow the algorithm to autonomously identify relevant information in a more complex and non-localized manner, as exemplified by neural networks like those used in CellCNN [43]. This approach bypasses the need for potentially biased decisions regarding how best to characterize marker intensity distributions (e.g., median, standard deviation) or the optimal selection of clusters representing cell subpopulations. Despite the reduced interpretability of this methods, recent advancements in deep learning architectures and their applications in automated diagnosis and monitoring present a promising avenue for extending these techniques to relapse prediction [55, 56, 57, 58].

To sum up, we have performed a machine learning-based relapse classification study involving 188 patients diagnosed with childhood BCP-ALL. A detailed characterization of immunophenotype and different cluster resolutions have been unable to distinguish relapse from non-relapse patients, and other algorithms from the literature exhibited similar outcomes. We conclude that different characterizations of FC data are required to uncover its potential prognostic value, pending the availability of high-dimensional omics data at diagnosis and more advanced cytometers that circumvent some of the challenges found throughout our study.

## 5. Supplemental Information

Supplemental information can be found online at XXXX.

## 6. CRediT authorship contribution statement

**Álvaro Martínez-Rubio**: Data curation, Formal analysis, Investigation, Methodology, Software, Writing—original draft, Writing—review & editing. **Salvador Chulián**: Data curation, Investigation, Writing—review & editing. **Ana Niño-López**: Data curation, Investigation, Writing—review & editing. **Rocío Picón-González**: Data curation, Investigation, Writing—review & editing. **Juan F. Rodríguez Gutiérrez**: Data curation, Resources, Writing—review & editing. **Eva Gálvez de la Villa**: Data curation, Resources, Writing—review & editing. **Teresa Caballero Velázquez**: Data curation, Resources, Writing—review & editing. **Águeda Molinos Quintana**: Data curation, Resources, Writing—review & editing. **Ana Castillo Robleda**: Data curation, Resources, Writing—review & editing. **Manuel Ramírez Orellana**: Data curation, Resources, Writing—review & editing. **María V. Martínez Sánchez**: Data curation, Resources, Writing—review & editing. **Alfredo Minguela Puras**: Data curation, Resources, Writing—review & editing. **José L. Fuster Soler**: Data curation, Resources, Writing—review & editing. **Cristina Blázquez Goñi**: Conceptualization, Data curation, Project administration, Resources, Writing—review & editing. **Víctor M. Pérez García**: Conceptualization, Funding acquisition, Project administration, Supervision, Writing—review & editing. **María Rosa**: Conceptualization, Funding acquisition, Project administration, Supervision, Writing—review & editing.

## 7. Declaration of competing interests

The authors declare no conflicts of interest.

## Acknowledgments

This work was partially supported by project PDC2022-133520-I00 funded by Ministerio de Ciencia e Innovación/ Agencia Estatal de investigación (doi:10.13039/501100011033) and European Union NextGenerationEU/PRTR; by project PID2022-140451OA-I00 funded by Ministerio de Ciencia e Innovación/Agencia Estatal de investigación (doi:10.13039/501100011033) and ERDF A way of making Europe; and by University of Castilla-La Mancha / ERDF, A way of making Europe (Applied Research Projects) under grant 2022-GRIN-34405. The support of Fundación Española para la Ciencia y la Tecnología (FECYT project PR214), Asociación Pablo Ugarte (APU, Spain) and Junta de Andalucía (Spain) group FQM-201 is also acknowledged. This work was also subsidized in its early stages by a grant for the research and biomedical innovation in the health sciences within the framework of the Integrated Territorial Initiative (ITI) for the province of Cádiz (grant number ITI-0038-2019).

## 8. Generative AI usage

During the preparation of this work, the authors used chatGPT (powered by OpenAI’s language model, GPT-3.5; http://openai.com) in order to improve readability and language of the work. After using this tool, the authors reviewed and edited the content as needed and take full responsibility for the content of the published article.

## Notes

### Competing Interest Statement

The authors have declared no competing interest.

### Funding Statement

This work was partially supported by project PDC2022-133520-I00 funded by Ministerio de Ciencia e Innovaci&oacuten/ Agencia Estatal de investigaci&oacuten (doi:10.13039/501100011033) and European Union NextGenerationEU/PRTR; by project PID2022-140451OA-I00 funded by Ministerio de Ciencia e Innovaci&oacuten/Agencia Estatal de investigaci&oacuten (doi:10.13039/501100011033) and ERDF A way of making Europe; and by University of Castilla-La Mancha / ERDF, A way of making Europe (Applied Research Projects) under grant 2022-GRIN-34405. The support of Fundaci&oacuten Espa&ntildeola para la Ciencia y la Tecnolog&iacutea (FECYT project PR214), Asociaci&oacuten Pablo Ugarte (APU, Spain) and Junta de Andaluc&iacutea (Spain) group FQM-201 is also acknowledged. This work was also subsidized in its early stages by a grant for the research and biomedical innovation in the health sciences within the framework of the Integrated Territorial Initiative (ITI) for the province of C&aacutediz (grant number ITI-0038-2019).

### Author Declarations

IRB of Hospital Universitario de Jerez de la Frontera gave ethical approval for this work. IRB of Hospital Infantil Universitario Niño Jesús gave ethical approval for this work. IRB of Hospital Universitario Virgen del Rocío gave ethical approval for this work. IRB of Hospital Clínico Universitario Virgen de la Arrixaca gave ethical approval for this work.

### Summary of Updates

Methodology has been updated, as well as result figures. Conclusions remains largely the same as the initial version.

