## Supplementary Material for "Computational flow cytometry immunophenotyping at diagnosis is unable to predict relapse in childhood B-cell Acute Lymphoblastic Leukemia"

| | Tubes | PerCP -Cy5-5-A | Pacific -Blue-A * | PE-Cy7 -A | PE-A | FITC-A | AmCyan -A ** | APC-Cy7 -A | APC-A |
|---|---|---|---|---|---|---|---|---|---|
| HNJ | Tube 1 | CD34 | CD3 | CD19 | CD79A | MPO | CD45 | CD3c | CD7 |
| | Tube 2 | CD13 | KAPPA | CD19 | IgMc | TdT | CD45 | LAMBDA | CD22c |
| | Tube 3 | CD34 | CD9 | CD19 | NG2 | CD15 | CD45 | CD24 | CD22 |
| | Tube 4 | CD34 | HLADR | CD19 | IgMs | CD2 | CD45 | - | CD33 |
| | Tube 5 | CD34 | - | CD19 | TSLPR | CD81 | CD45 | CD38 | CD52 |
| | Tube 6 | CD34 | CD20 | CD19 | CD66c | CD58 | CD45 | CD38 | CD10 |
| HVA | Tube 1 | CD38 | CD4 | CD19 | CD79B | CD15 | CD3 | CD45 | CD14+CD8 |
| | Tube 2 | HLADR | CD19 | CD34 | CD13 | CD66c | - | CD45 | CD33 |
| | Tube 3 | CD58 | CD20 | CD10 | IgMs | CD19 | - | CD45 | CD22 |
| | Tube 4 | CD10 | CD20 | CD19 | IgMc | CD79Ac | - | CD45 | CD34 |
| | Tube 5 | CD10 | CD3c | CD19 | MPO | TdT | - | CD45 | CD34 |
| HVR | Tube 1 | CD34 | CD21 | CD19 | NG2 | CD15+CD65 | CD45 | CD81 | CD123 |
| | Tube 2 | CD34 | CD20 | CD19 | CD66c | CD58 | CD45 | CD38 | CD10 |
| | Tube 3 | CD34 | KAPPA | CD19 | CD33 | IgM | CD45 | LAMBDA | IgM+CD117 |
| | Tube 4 | CD34 | CD9 | CD19 | CD13 | TdT | CD45 | CD24 | CD22 |
| | Tube 5 | CD34 | CD3c | CD19 | CD79Ac | MPO | CD45 | CD3 | CD7 |

**Table S1:** Monoclonal fluorochrome-conjugated antibody combinations employed at hospitals HNJ, HVA, and HVR. * V450-A in HVR. ** V500-A in HVR.

| Algorithm | Year | Feature Extraction | Classification | Ref. |
|---|---|---|---|---|
| **Citrus** | 2014 | Cluster + MFI/Abund | Lasso regularized LR | [2] |
| **CellCNN** | 2017 | Convolutional Neural Network | | [1] |
| **Cydar** | 2017 | Cluster + MFI/Abund | GLM | [9] |
| **DDPR** | 2018 | Cluster + MFI/Abund+clinical | GLM | [5] |
| **Diffcyt** | 2019 | Cluster + MFI/Abund | GLM | [10] |
| **CytoDX** | 2019 | - | LR | [7] |
| **Fisher's Ratio** | 2020 | Percentile Vectors | Fisher's Ratio-based classifier | [3] |
| **DeepCNN** | 2020 | Deep Convolutional Neural Network | | [8] |
| **FAUST** | 2021 | Cluster+Abund | GLM | [6] |
| **CytoSet** | 2021 | Permutation Invariant Network (deep sets) | | [11] |
| **TDA** | 2023 | PH barcode summary + PI | RF, LR, SVM | [4] |

**Table S2: Selection of flow cytometry-based classification algorithms from the literature**. MFI = Median Fluorescence Intensity. PH = Persistent Homology. PI = Persistence Image. LR= Logistic Regression. GLM = Generalized Linear Model. RF = Random Forest. SVM = Support Vector Machine.

| Model | caret function | Preprocess | Hyperparameter | Values |
|---|---|---|---|---|
| KNN | knn | Center and scale | k | 6 integers between $0.2 \cdot n_{features}$ and $0.8 \cdot n_{features}$ |
| NB | naive_bayes | None | laplace | 1 |
| | | | usekernel | TRUE, FALSE |
| | | | adjust | 0.01, 0.1, 1 |
| RF | rf | None | mtry | 6 integers between $0.2 \cdot n_{features}$ and $0.8 \cdot n_{features}$ |
| SVM | svmLinearWeights | Center and scale | cost | 1, 10 |
| | | | weight | 0.01, 0.1, 1 |

**Table S3: Model hyperparameters for inner loop tuning.** KNN = K-Nearest Neighbors. NB = Naive Bayes Classifier. RF = Random Forest. SVM = Support Vector Machine. All models are included in the R package caret. $n_{features}$ equals the number of features present in each dataset.

| | Dataset 1 (HVR) (N=56) | Dataset 2 (HVA) (N=80) | Dataset 3 (HNJ) (N=116) | Total (N=252) |
|---|---|---|---|---|
| Sex - no. (%) | | | | |
| Male | 33 (58.9) | 42 (52.5) | 56 (48.3) | 131 (52.0) |
| Female | 23 (41.1) | 38 (47.5) | 60 (51.7) | 121 (48.0) |
| Age at diagnosis - yr | | | | |
| Median | 3 | 5 | 4 | 4 |
| Range | 0 - 13 | 0 - 15 | 0 - 16 | 0 - 16 |
| Long term status -no. (%) | | | | |
| Relapse | 12 (21.4) | 7 (8.8) | 25 (21.6) | 44 (17.5) |
| No relapse | 43 (76.8) | 73 (91.2) | 91 (78.4) | 207 (82.1) |
| Immunophenotype - no. (%) | | | | |
| Common | 36 (64.3) | 67 (83.8) | 108 (93.1) | 211 (83.7) |
| Pre-B | 16 (28.6) | 10 (12.5) | 5 (4.3) | 31 (12.3) |
| Pro-B | 3 (5.4) | 3 (3.8) | 3 (2.6) | 9 (3.6) |
| Mixed | 1 (1.8) | 0 (0) | 0 (0) | 1 (0.4) |
| Bone Marrow blasts at diagnosis - % | | | | |
| Median | 80.4 | 77.0 | 84.0 | 80.9 |
| Range | 10.0 - 97.6 | 18.3 - 99.0 | 3.0 - 99.0 | 3.0 - 99.0 |
| Leukocytes - cell/nL | | | | |
| Median | 7.76 | 7.39 | 10.4 | 8.59 |
| Range | 0.6 - 494.0 | 0.54 - 336.19 | 0.21 - 294.0 | 0.21 - 336.19 |
| Central Nervous System involvement - no. (%) | | | | |
| Yes | 2 (3.6) | 3 (3.8) | 11 (9.5) | 16 (6.3) |
| No | 53 (94.6) | 77 (96.2) | 105 (90.5) | 235 (93.3) |
| Risk at diagnosis - no. (%) | | | | |
| High | 2 (3.6) | 6 (7.5) | 2 (1.7) | 10 (4.0) |
| Intermediate | 23 (41.1) | 47 (58.8) | 87 (75.0) | 157 (62.3) |
| Low | 31 (55.3) | 27 (33.7) | 27 (23.3) | 85 (33.7) |
| Karyotype - no. (%) | | | | |
| High hyperdiploidy (>50) | 13 (23.2) | 2 (2.5) | 15 (13.0) | 30 (12.0) |
| Hyperdiploidy (47-50) | 3 (5.4) | 2 (2.5) | 9 (7.7) | 14 (5.6) |
| Normal (46) | 22 (39.3) | 11 (13.8) | 56 (48.3) | 89 (35.3) |
| Hypodiploidy (40-45) | 2 (3.6) | 0 (0) | 5 (4.3) | 7 (2.7) |
| Low hypodiploidy (<40) | 1 (1.8) | 0 (0) | 0 (0) | 1 (0.4) |
| No metaphases | 14 (25.0) | 9 (11.2) | 28 (24.1) | 51 (20.2) |
| No information | 1 (1.8) | 56 (70.0) | 3 (2.6) | 60 (23.8) |
| Chromosomic alterations - no. (%) | | | | |
| ETV6/RUNX1 t(12;21) | 11 (19.6) | 18 (22.5) | 29 (25.0) | 58 (23.0) |
| TCF3/PBX1 t(1;19) | 2 (3.6) | 1 (1.2) | 5 (4.3) | 8 (3.2) |
| MLL rearrangement | 5 (8.9) | 3 (3.8) | 2 (1.7) | 10 (4.0) |
| BCR/ABL1 t(9;22) | 0 (0) | 1 (1.2) | 3 (2.6) | 4 (1.6) |
| No alterations | 36 (64.3) | 53 (66.3) | 76 (65.5) | 165 (65.4) |
| No information | 2 (3.6) | 4 (5.0) | 1 (0.8) | 7 (2.8) |

**Table S4:** Summary of the clinicopathologic characteristics of the full cohort.

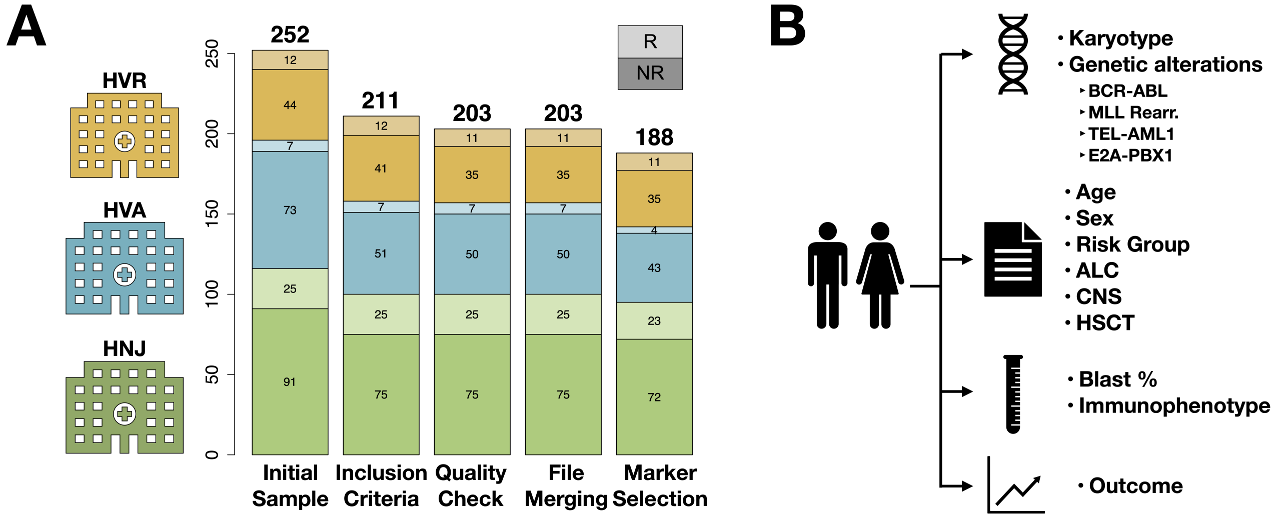


**Figure S1:** **A.** Number of patients retained for analysis at each preprocessing step. See 'Methods' section for a detailed description of each step. Color denotes hospital. **B.** Source and type of data collected for the study.

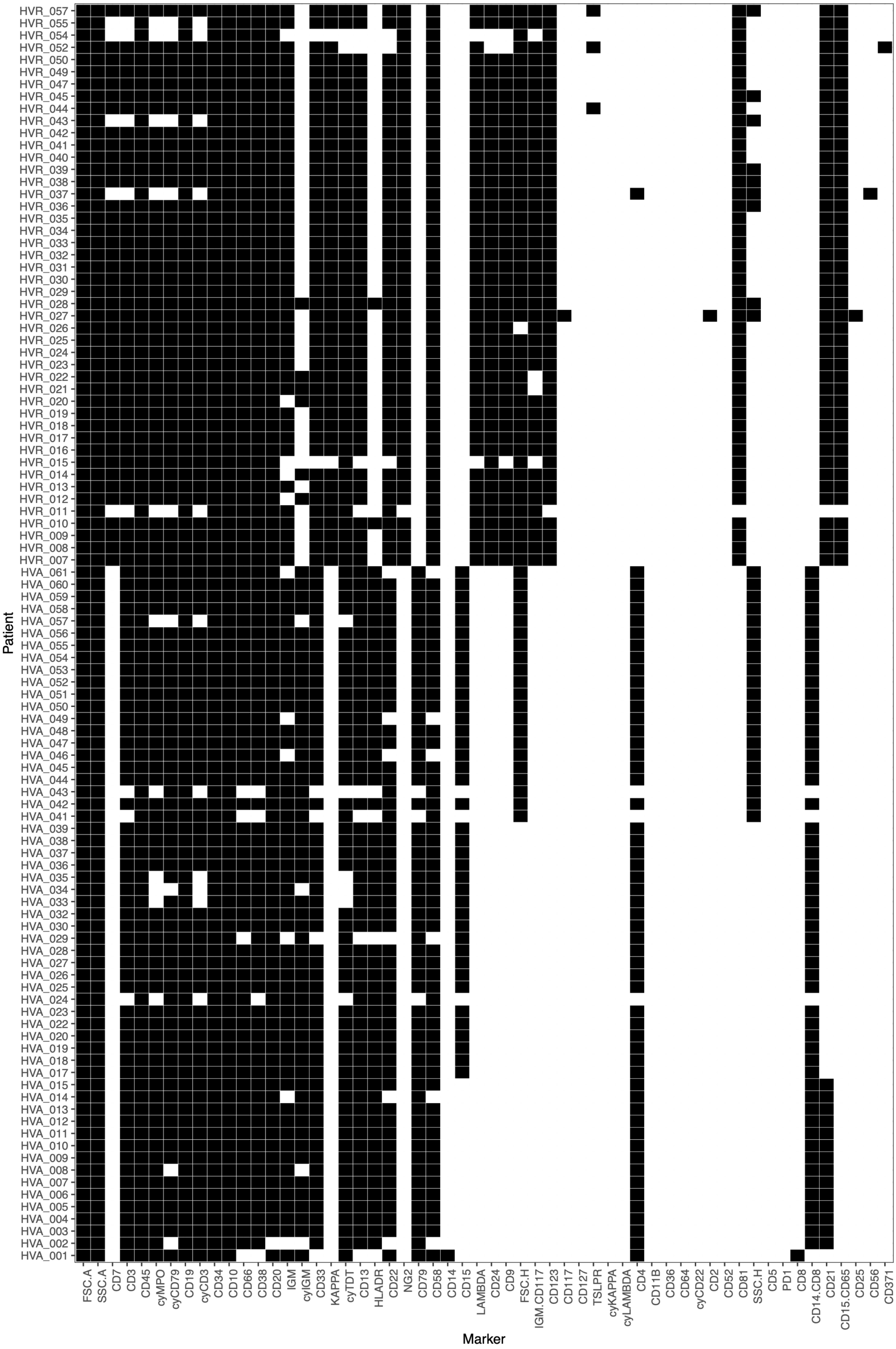


**Figure S2:** Markers per patient in hospitals HVA and HVR

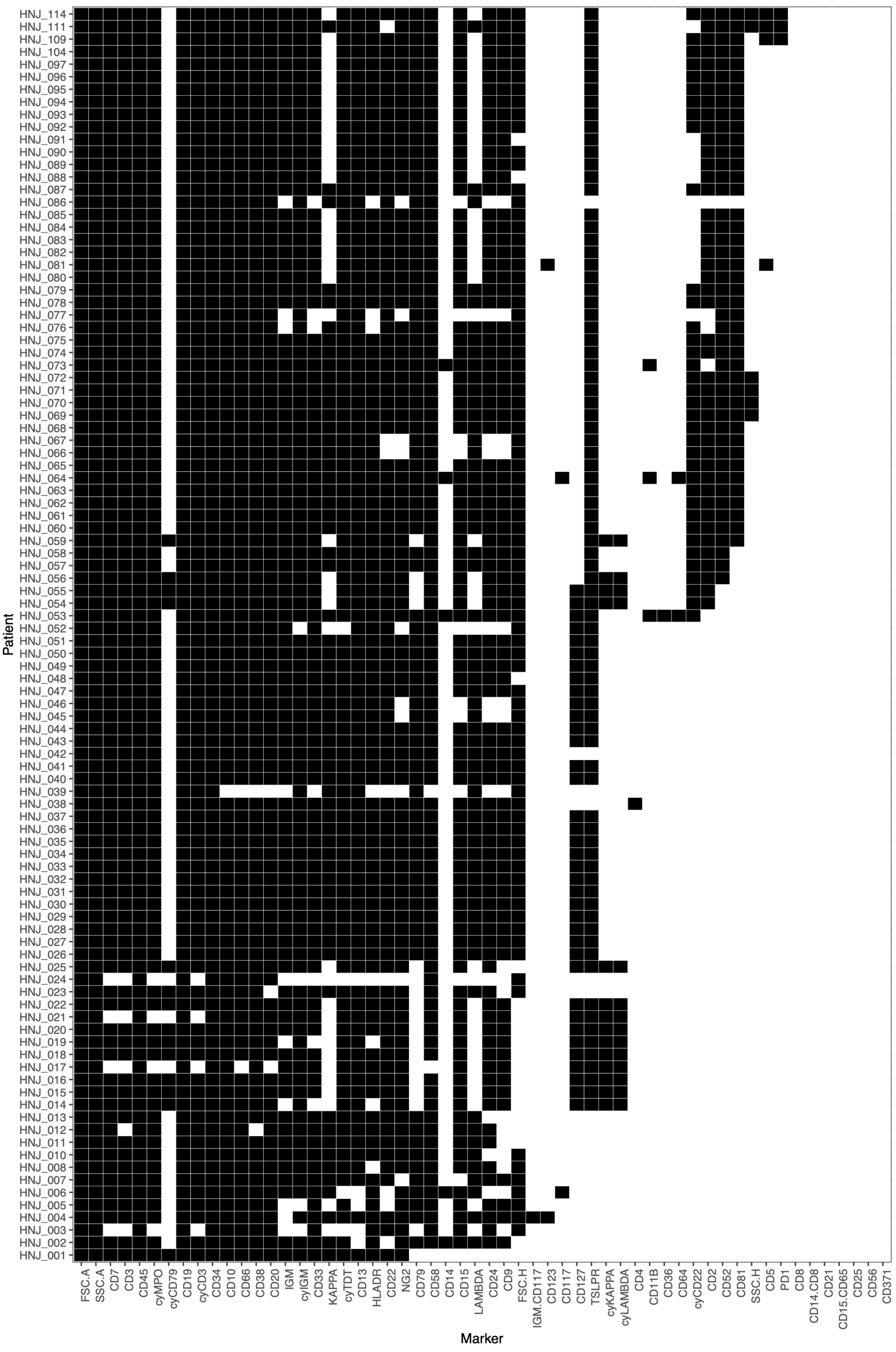


**Figure S3:** Markers per patient in hospital HNJ

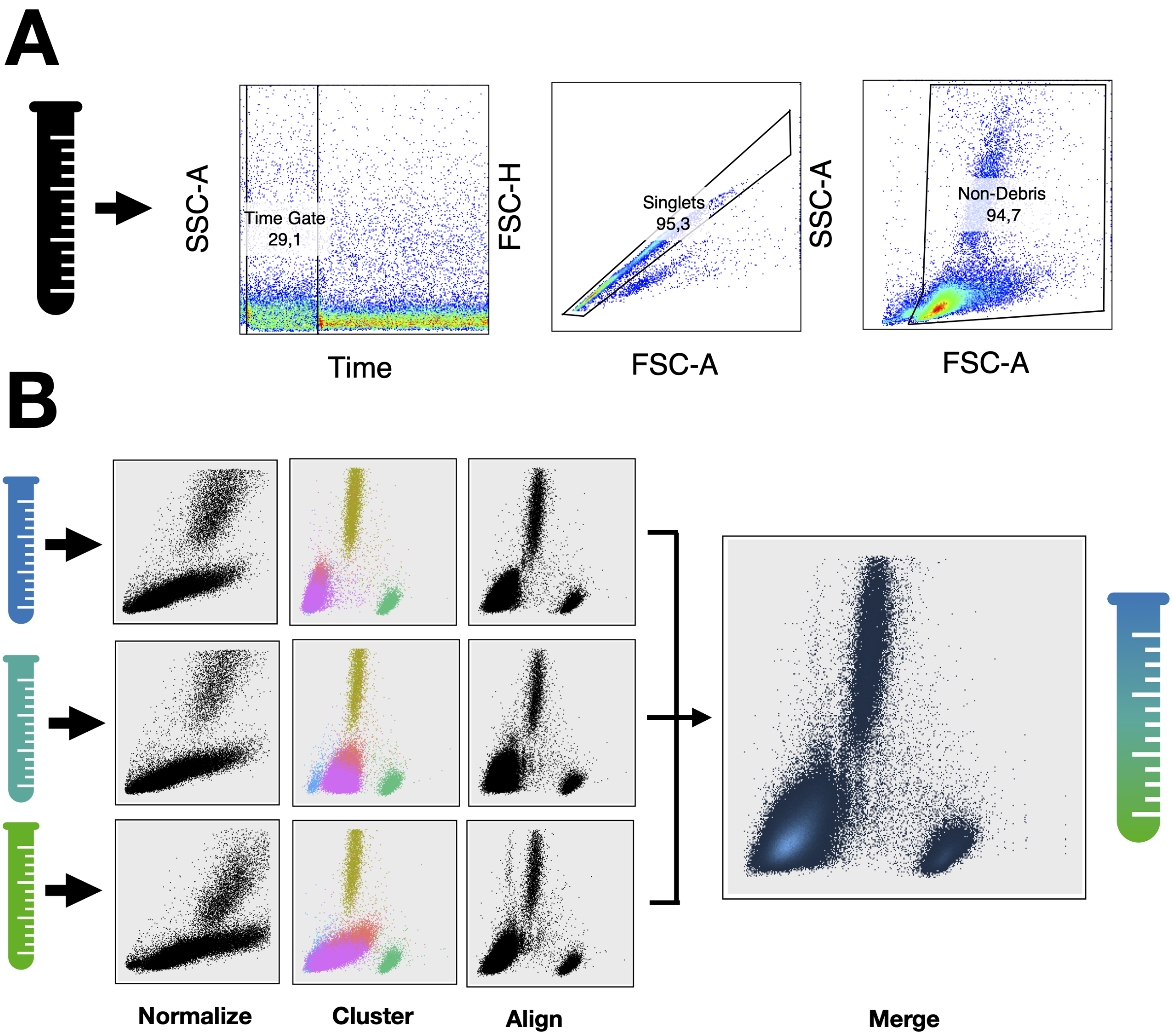


**Figure S4: Flow cytometry data preprocessing.** **A.** Manual step: Acquisition errors (Time-SSC.A), doublets (FSC.A-FSC.H) and debris (FSC.A-SSC.A) are gated out in FlowJo. **B.** Computational step: Modified min-max normalization, quantile normalization and imputation are carried out in R.

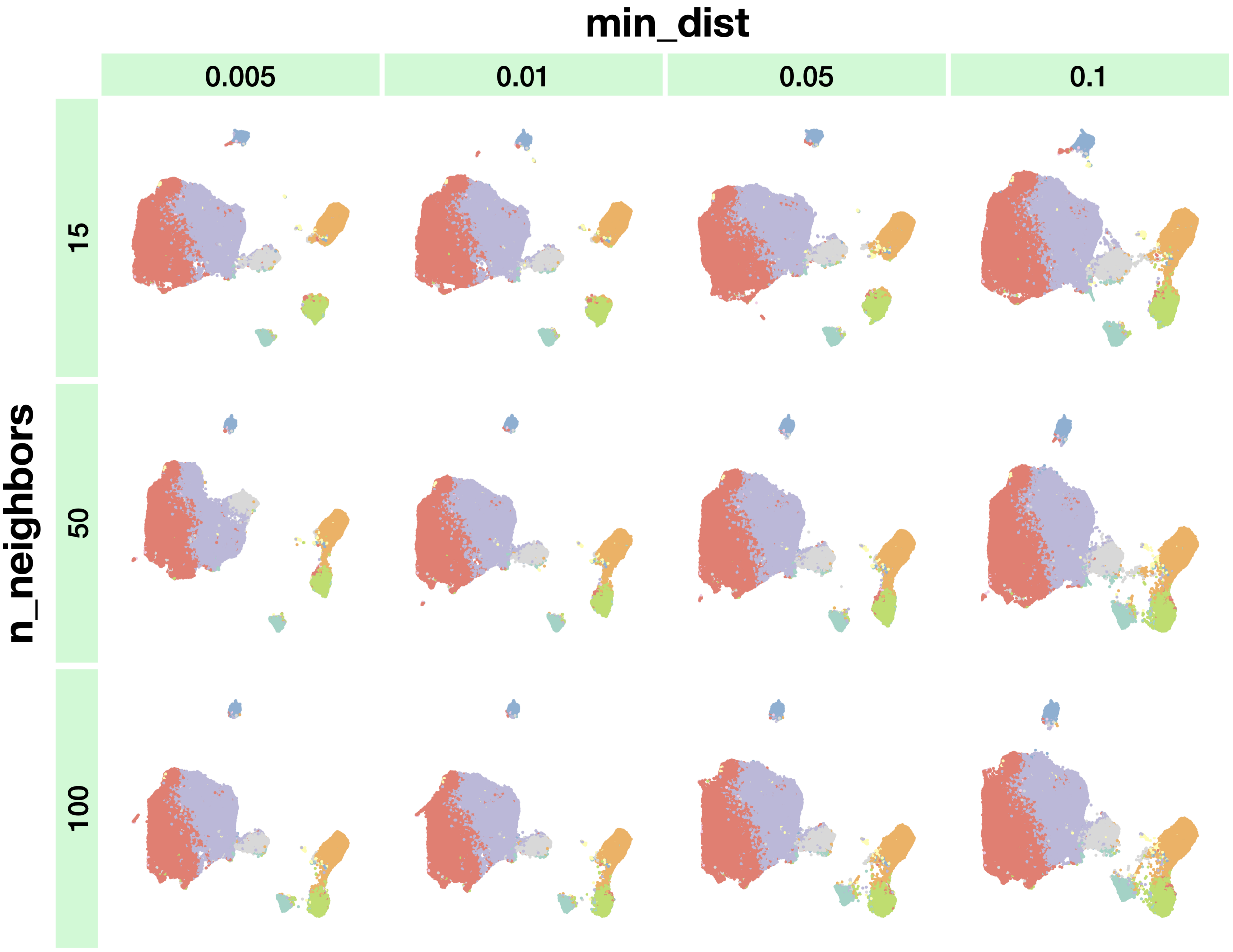


**Figure S5: UMAP hyperparameter exploration.** UMAP embedding of the cohort of 188 patients for different values of UMAP hyperparameters $min\_dist$ and $n\_neighbors$. The variations are not very significant. For larger values of the hyperparameters the embedding loses local structure and thus the metaclusters become too close. The final selection was $min\_dist = 0.01$ and $n\_neighbors = 15$

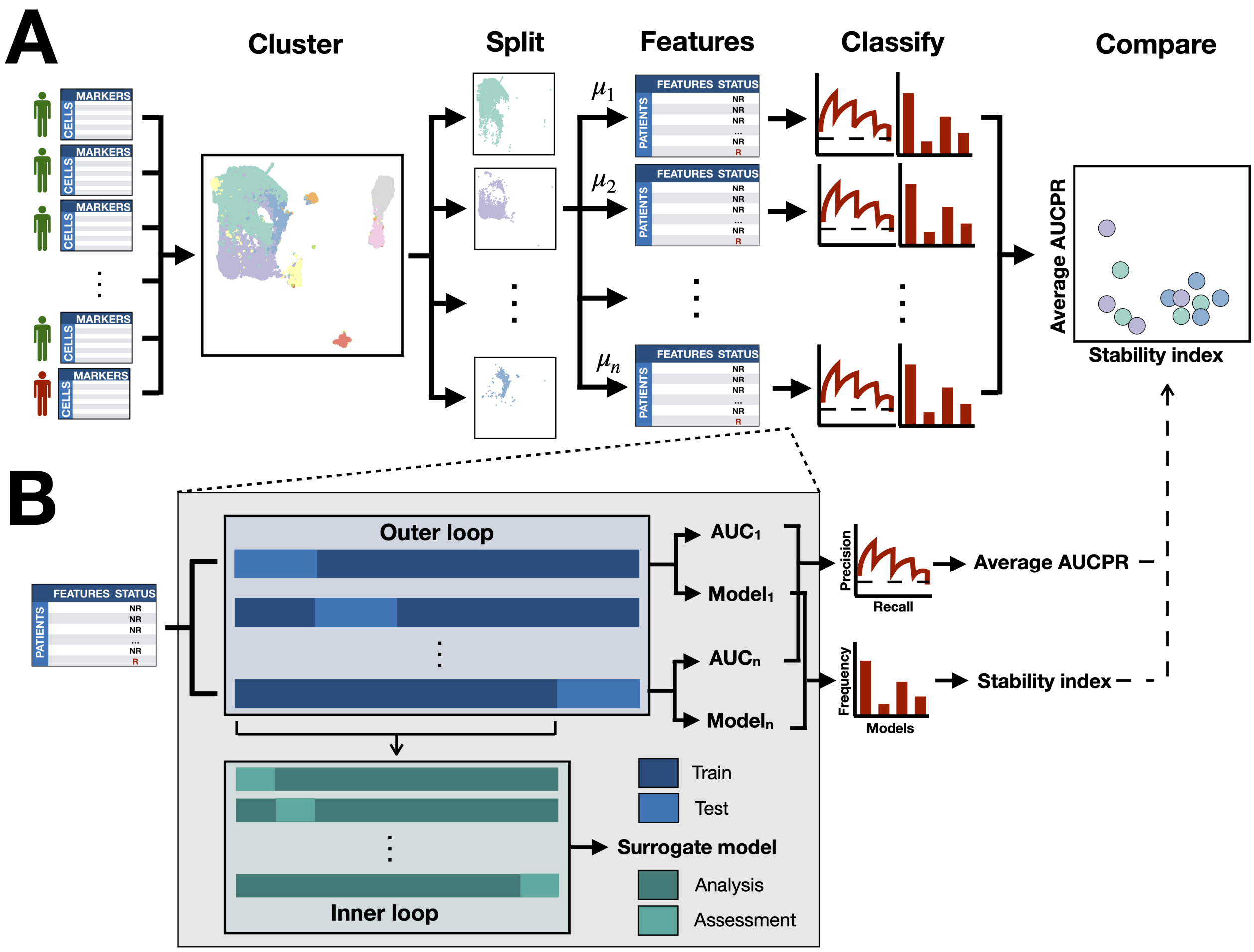


**Figure S6: Workflow for classification-based biomarker discovery. A.** Description of the steps that transform the flow cytometry database into metrics of performance for each feature and each metacluster. After pooling all patient data and clustering with FlowSOM, each metacluster is analysed separately (Split step). Marker expression is summarized by means of different statistical moments $\mu_i$. For each moment or metric, a dataset is constructed (Features step). Each dataset is fed to the classification routine, yielding an averaged Precision-Recall curve and a measure of the stability of the routine (Classify step). The results for all metaclusters and metrics are then visualized together (Compare step). **B.** Detailed view of the classification routine. In the datasets constructed, each row represents an individual (patients) and each column the statistical moment computed for each marker (features). The dataset is first split in an outer cross-validation loop that serves to estimate the performance of the classifier in unseen data. For each fold, an inner cross-validation loop is designed that serves to select the optimal algorithm among Naive Bayes, K-nearest neighbors, Random Forest and Linear Support Vector Machine. The result of this nested cross-validation scheme is an estimation of performance estimation (Average AUCPR) and a measure of the stability of the classification routine, which is derived from the frequencies with which these four algorithms are selected (Stability Index).

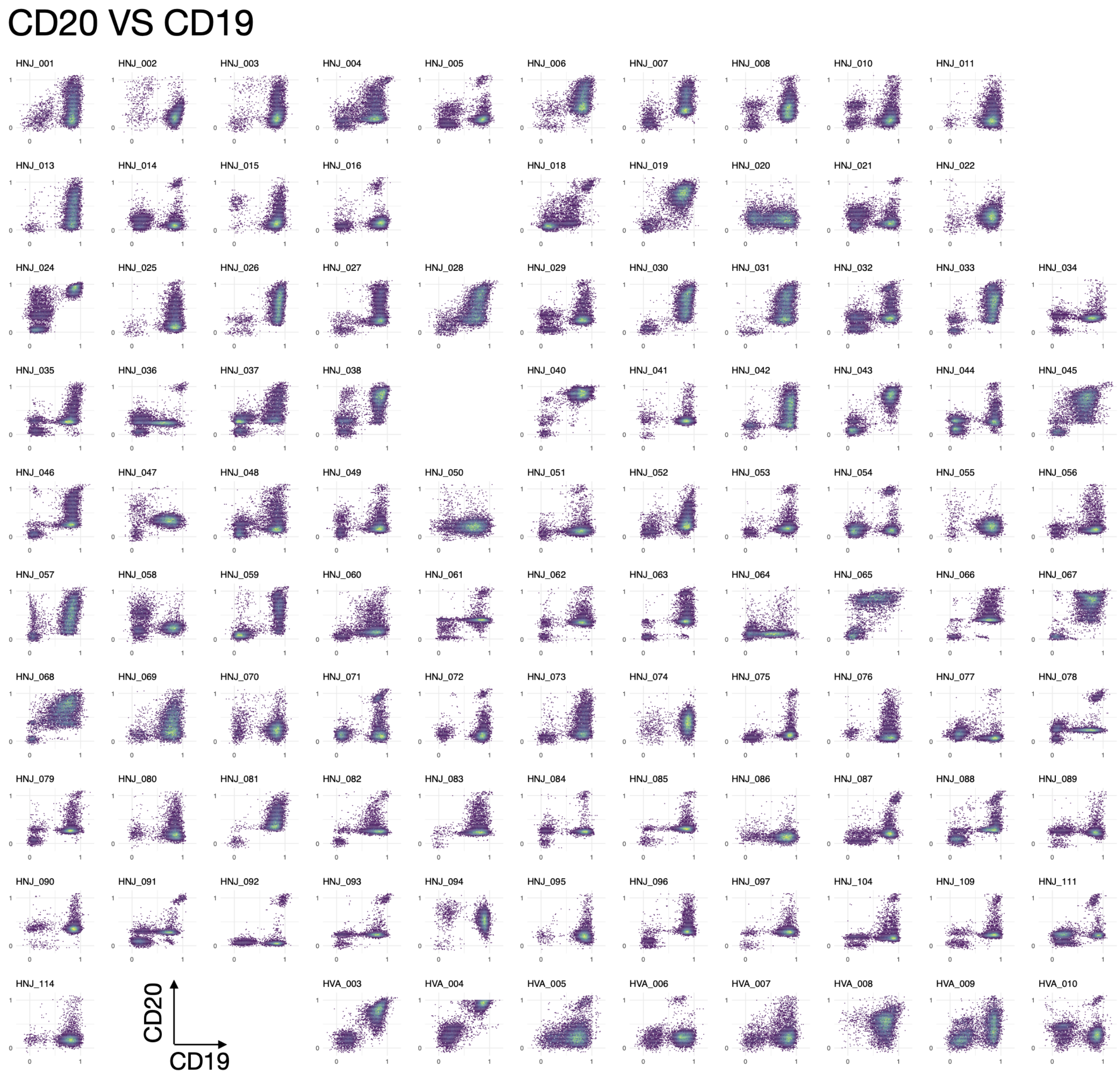


**Figure S7: Bidimensional representation of merged data (I).** Imputed marker CD20 vs backbone marker CD19.

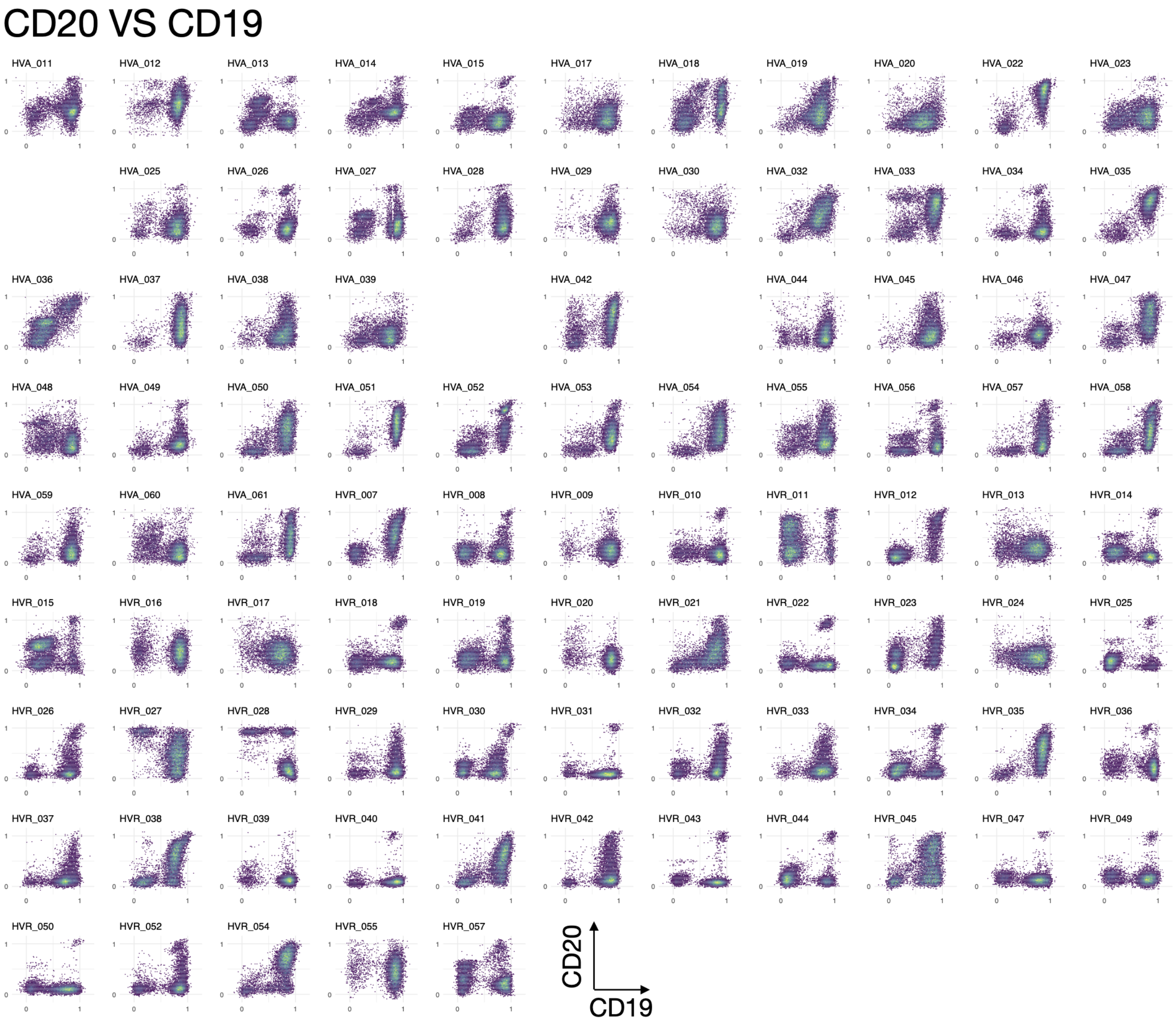


**Figure S8: Bidimensional representation of merged data (II).** Imputed marker CD20 vs backbone marker CD19.

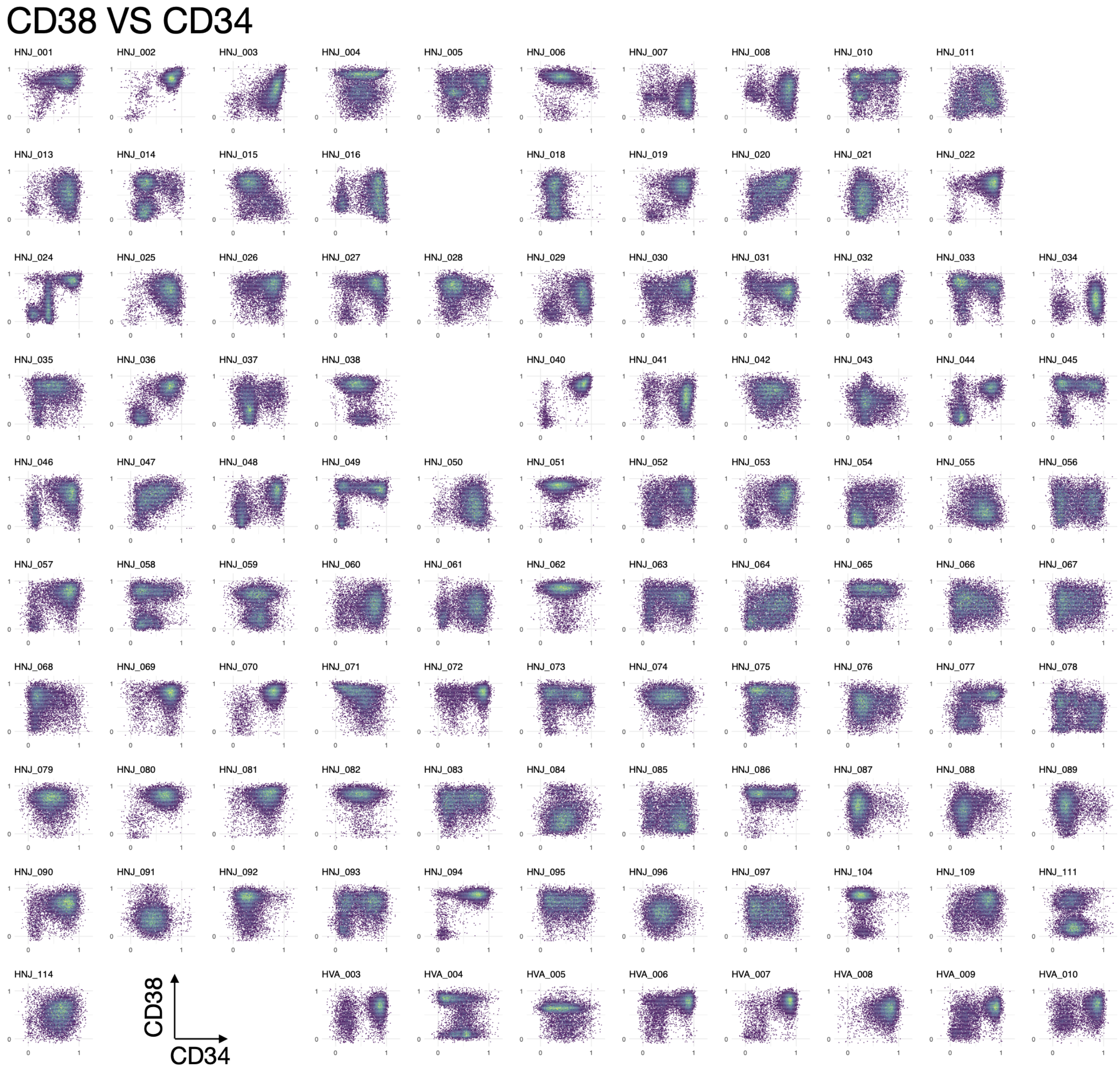


**Figure S9: Bidimensional representation of merged data (III).** Imputed marker CD38 vs backbone marker CD34.

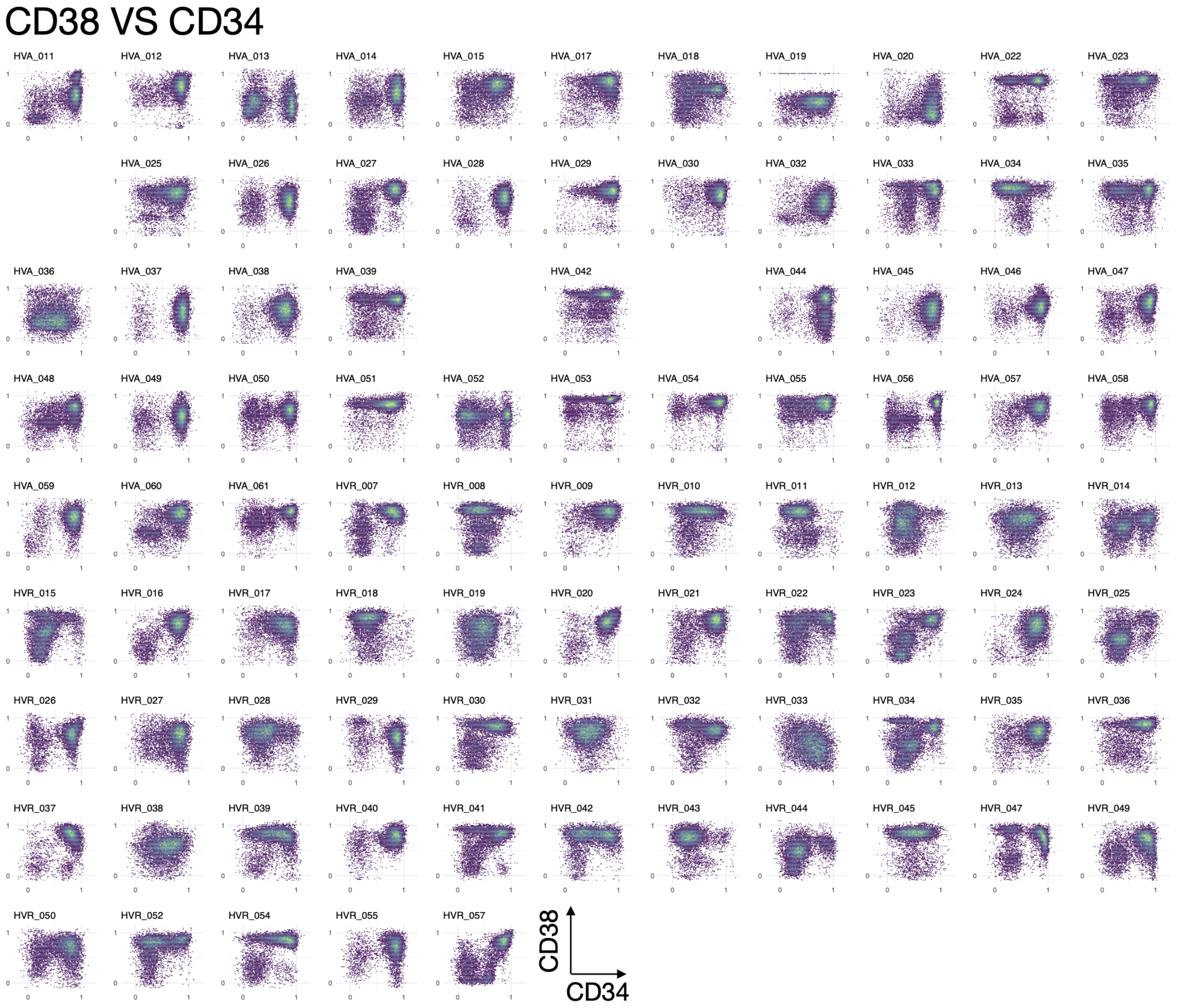


**Figure S10: Bidimensional representation of merged data (IV).** Imputed marker CD38 vs backbone marker CD34.

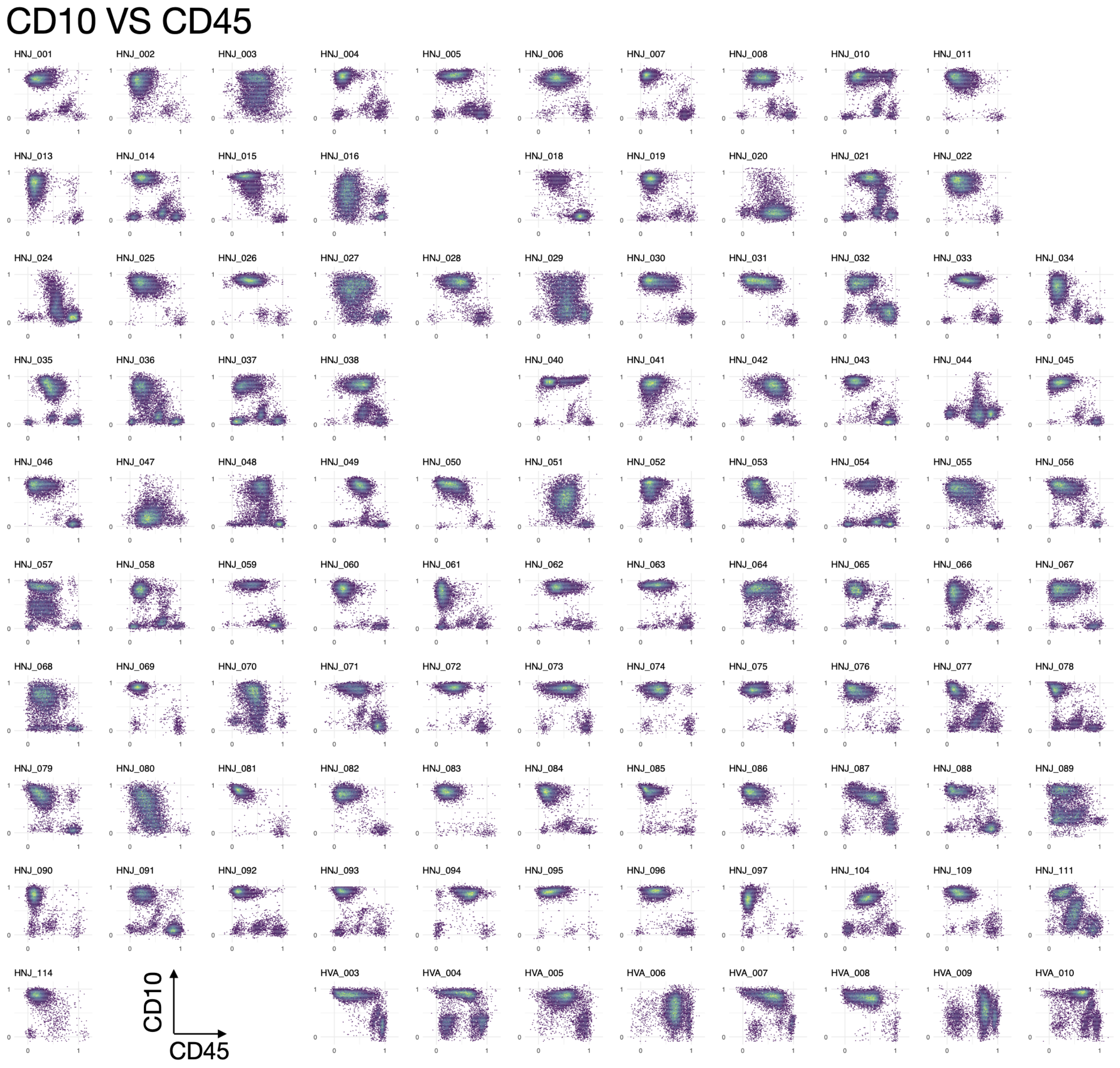


**Figure S11: Bidimensional representation of merged data (V).** Imputed marker CD10 vs backbone marker CD45.

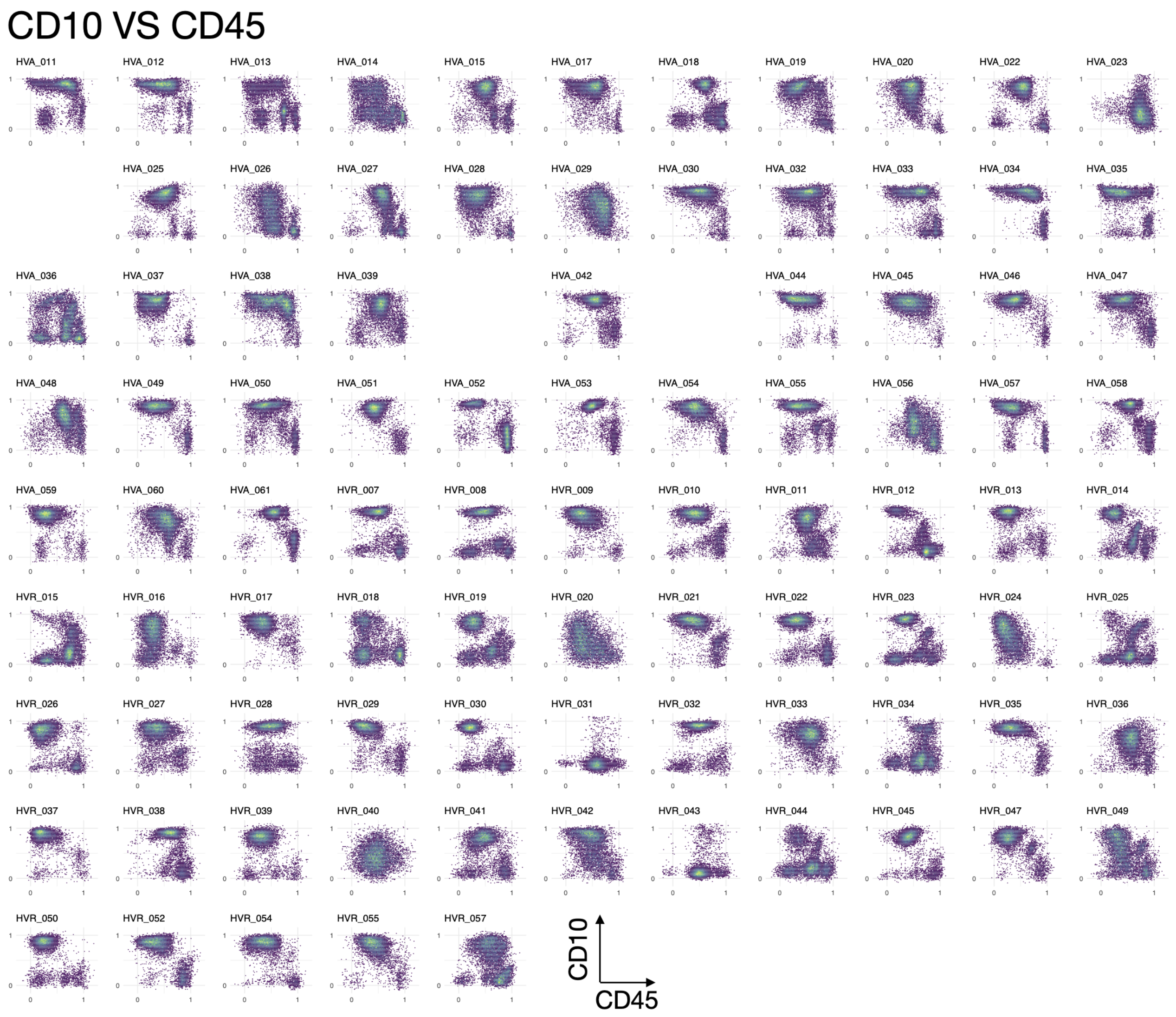


**Figure S12: Bidimensional representation of merged data (VI).** Imputed marker CD10 vs backbone marker CD45.

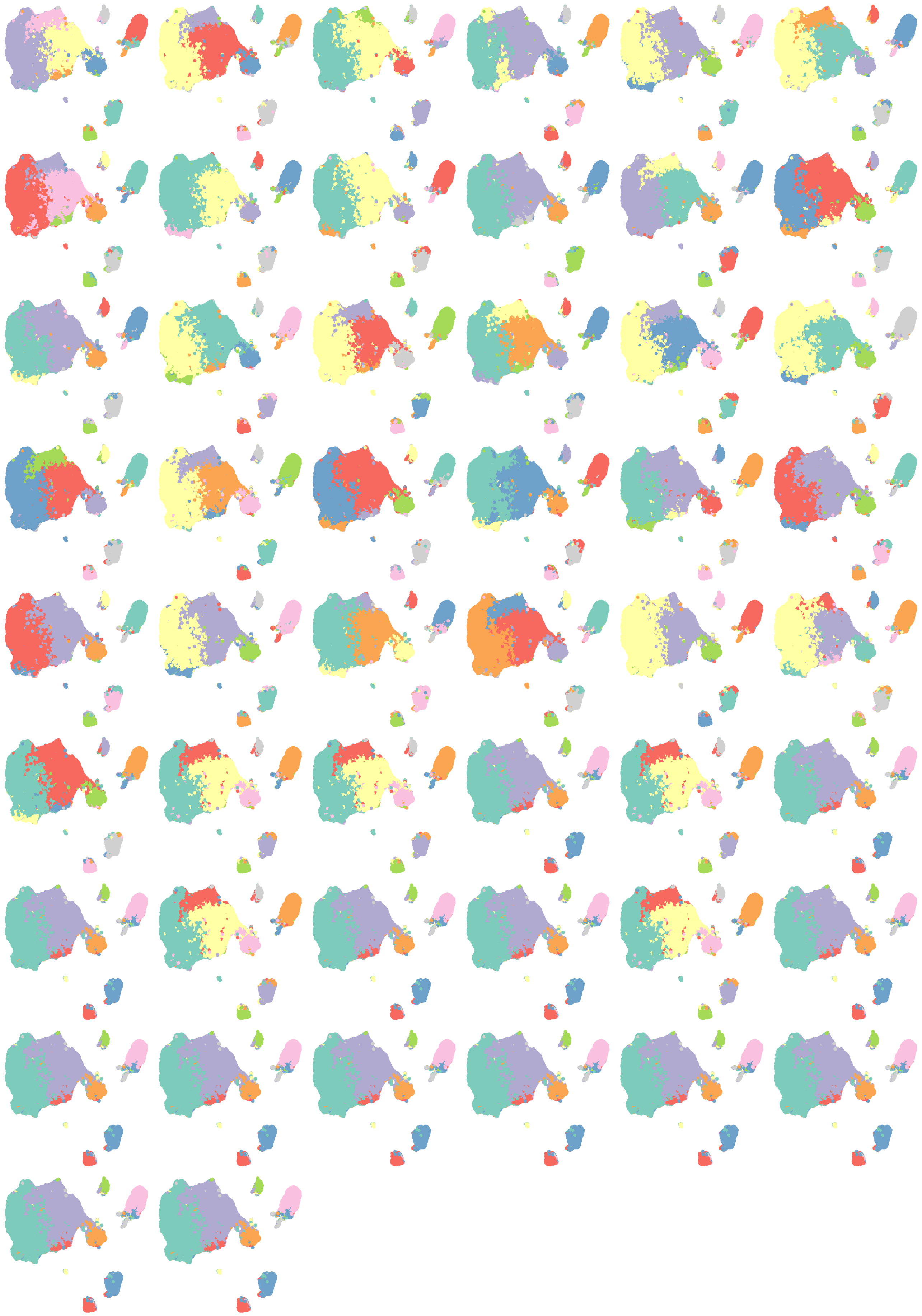

**Figure S13: Individual iterations of FlowSOM metaclustering for $k = 9$.** UMAP embedding with cell color given by metacluster, for 50 different iterations of the algorithm. The consensus of the 50 iterations was selected as the final metacluster assignment shown in Figure 2.

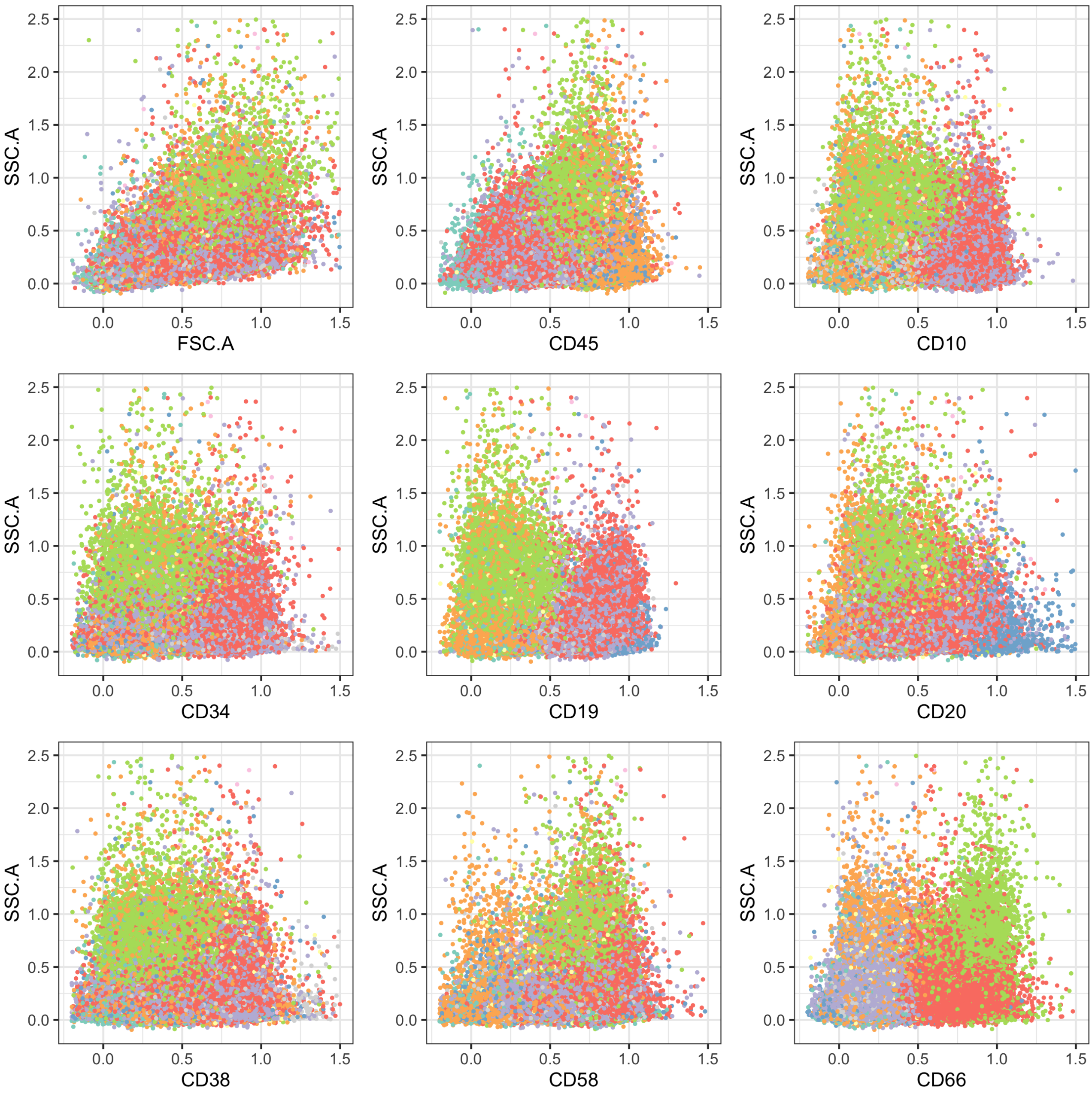


**Figure S14: Bidimensional representation of markers in patient selection A .** We compare marker expression with side-scatter. These classical visualizations help confirm the manual annotation performed on the UMAP embedding (Figure 2).

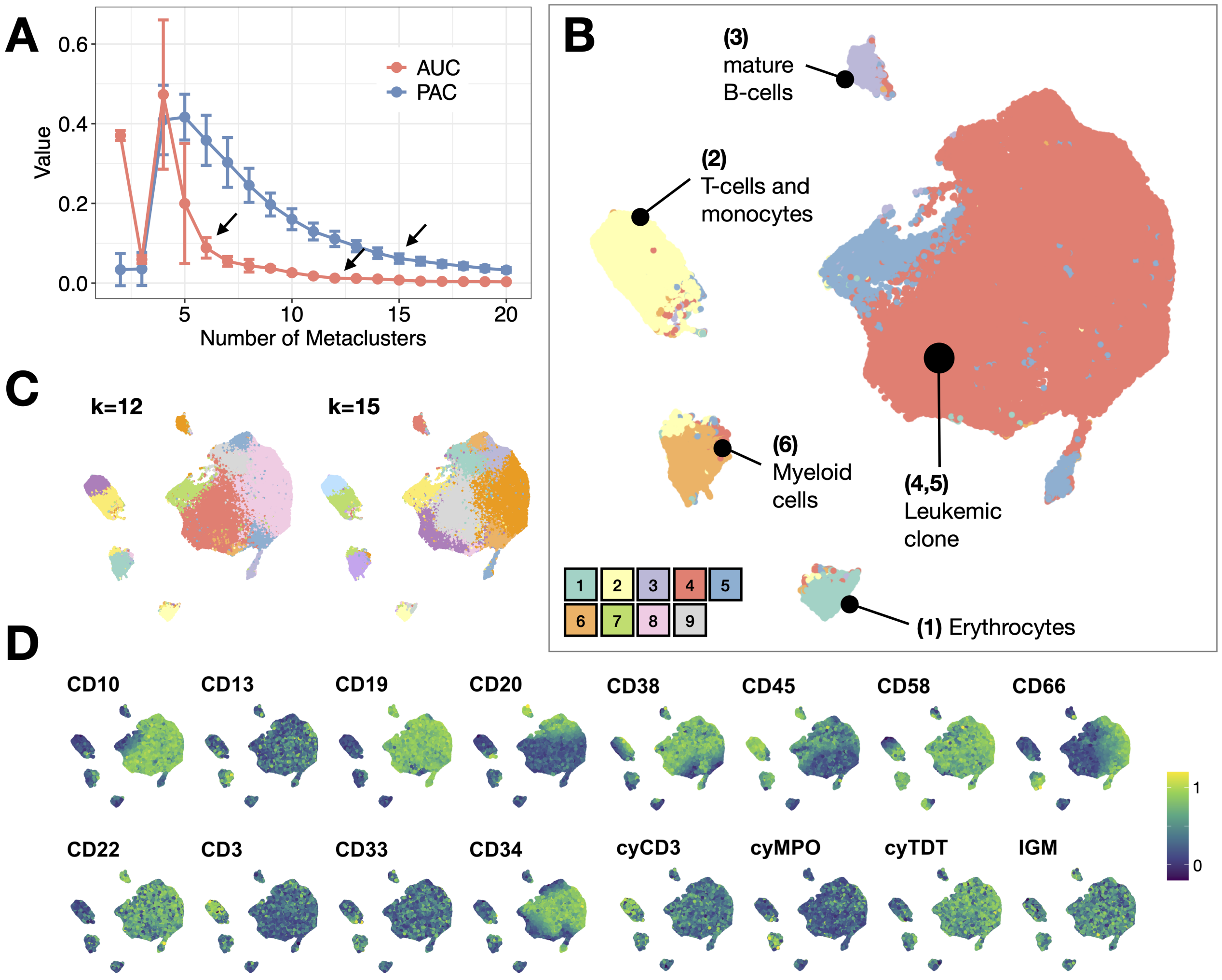


**Figure S15: Clustering and visualization of flow cytometry data for selection B.** **A.** Values of AUC (Area Under the Curve of the empirical distribution function of a consensus matrix) and PAC (Proportion of Ambiguous Clustering) according to number of metaclusters selected by FlowSOM algorithm in Selection A. **B.** UMAP visualization of 6 FlowSOM metaclusters in Selection B, manually labelled according to marker expression. **C.** UMAP visualization of 12 metaclusters and 15 metaclusters in Selection B. **D.** Relative intensity of marker expression in the UMAP embedding employed for visualization.

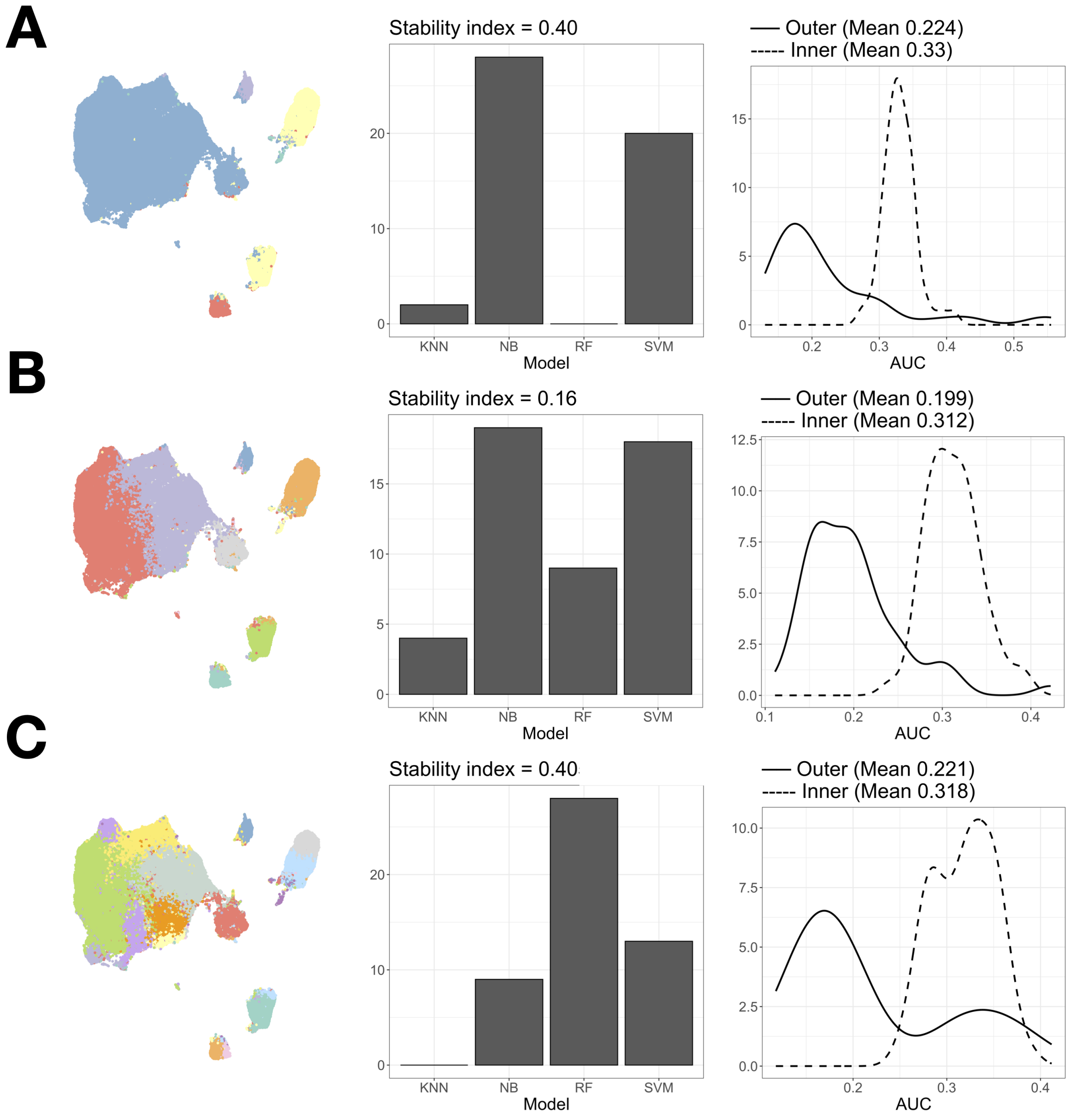


**Figure S16: Reliability assessment of abundance-based classification. A.** Classification results for 5 metaclusters. First panel shows the number of times each model is selected in the inner routine of the cross-validation scheme. More stable schemes tend to select the same model. This is quantified with the stability index (See 'Methods' in the main text). The next panel shows the comparison between the area under Precision-Recall curve for the outer and inner loops of the nested-cross validation scheme. Overfitted models would display strong differences between both distributions. **B.** Classification results for 9 metaclusters. **C.** Classification results for 15 metaclusters.

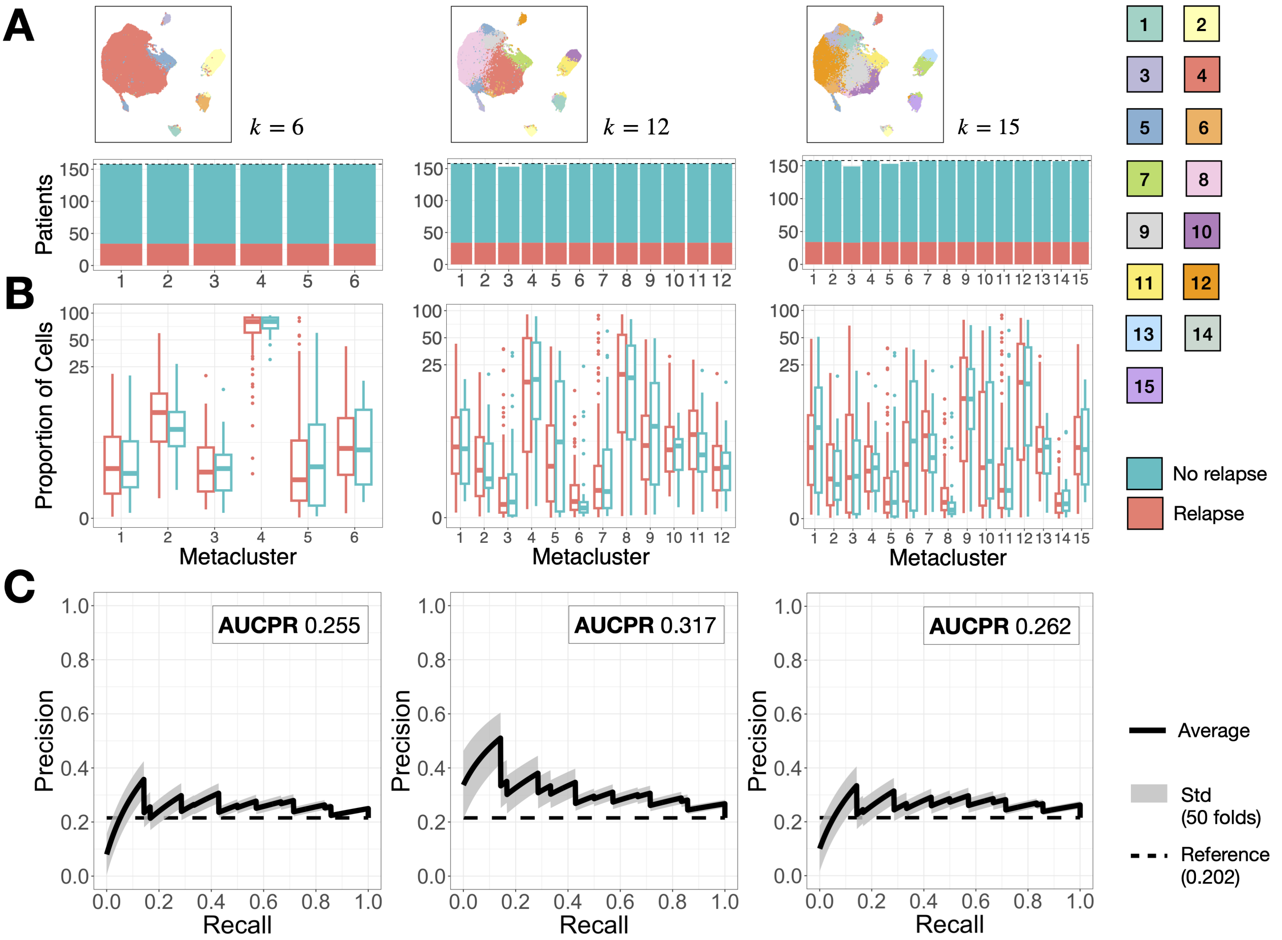


**Figure S17: Results of abundance-based classification for selection B. A.** Number of patients included in each metacluster, for the three selections of optimal number of metaclusters (Figure 2).**B.** Comparison of cell percentage per cluster between relapse (R) and non-relapse (NR) patients. Boxplot includes median and IQR. The scale has been transformed with an inverse hyperbolic sine for clarity. **B.** Classification results in terms of Area Under the Precision Recall Curve using information from all metaclusters. The shaded region represents the standard deviation of 50 repetitions of the classification routine (10 folds + 5 repeats). Horizontal dashed line represents the baseline precision, which equals the proportion of relapse patients in our dataset (Table 1).

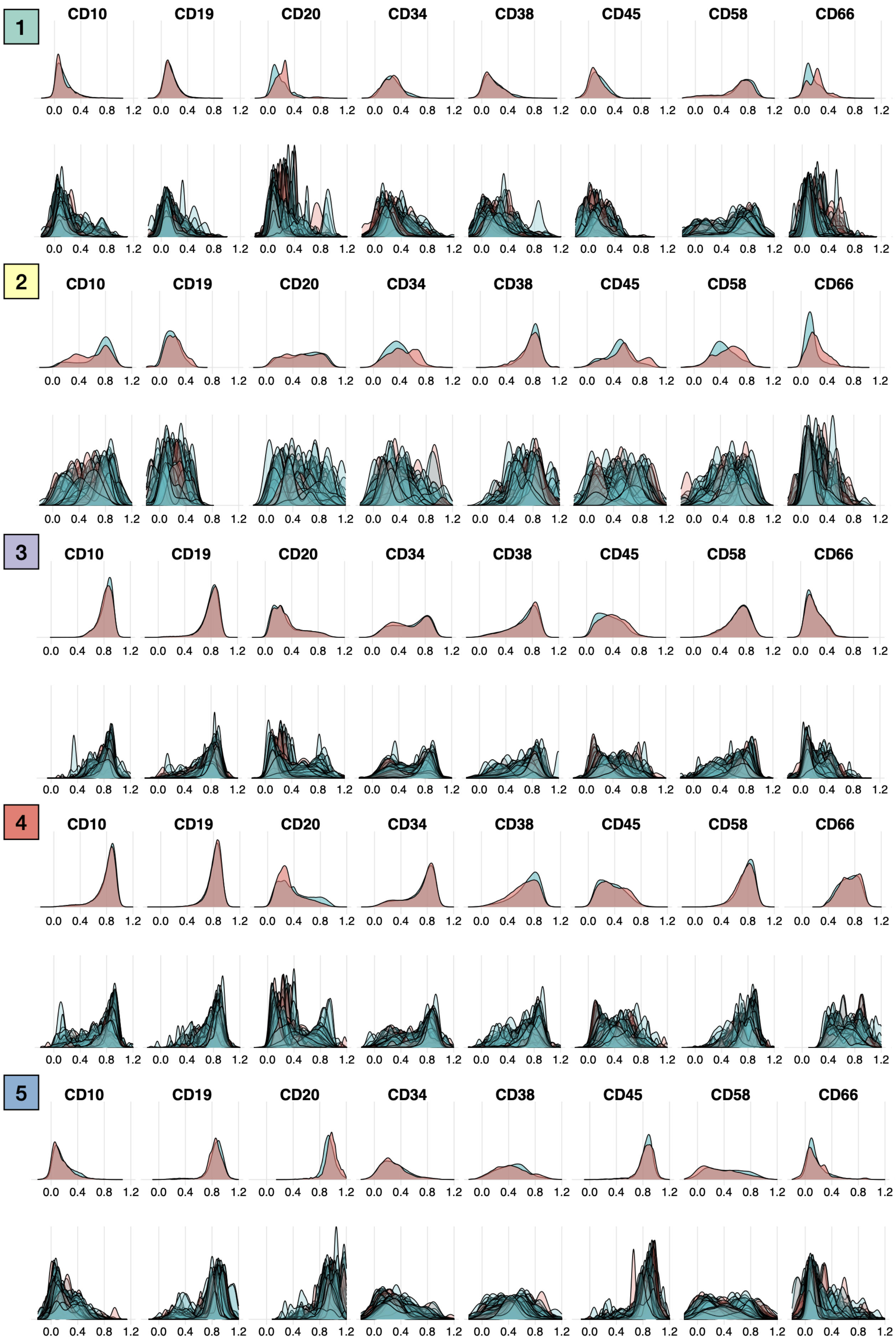


**Figure S18: Marker expression per metacluster (I).** Top row displays the intensity of expression considering all patients together. Bottom row segregates this information by patient.

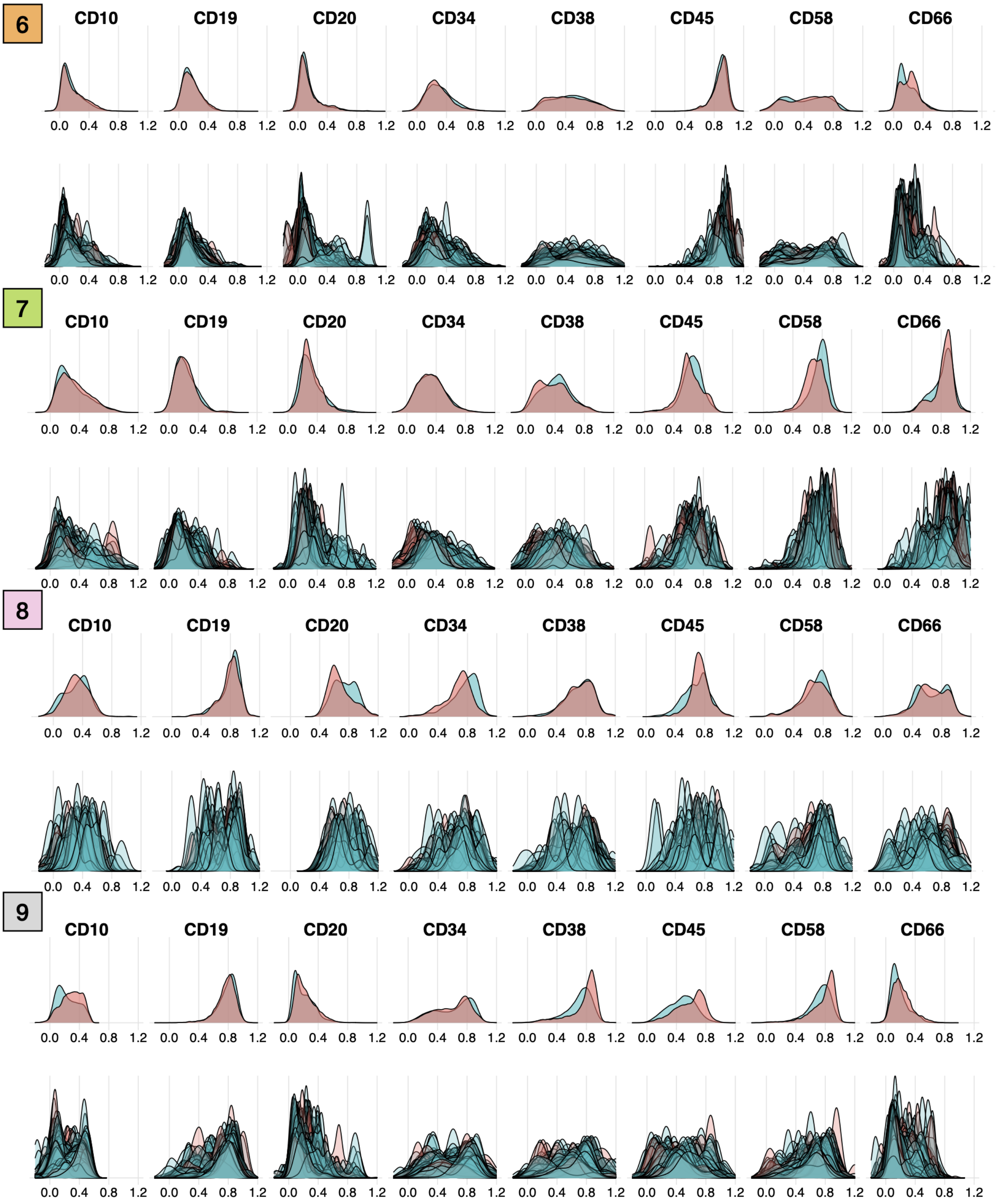


**Figure S19: Marker expression per metacluster (II).** Top row displays the intensity of expression considering all patients together. Bottom row segregates this information by patient.

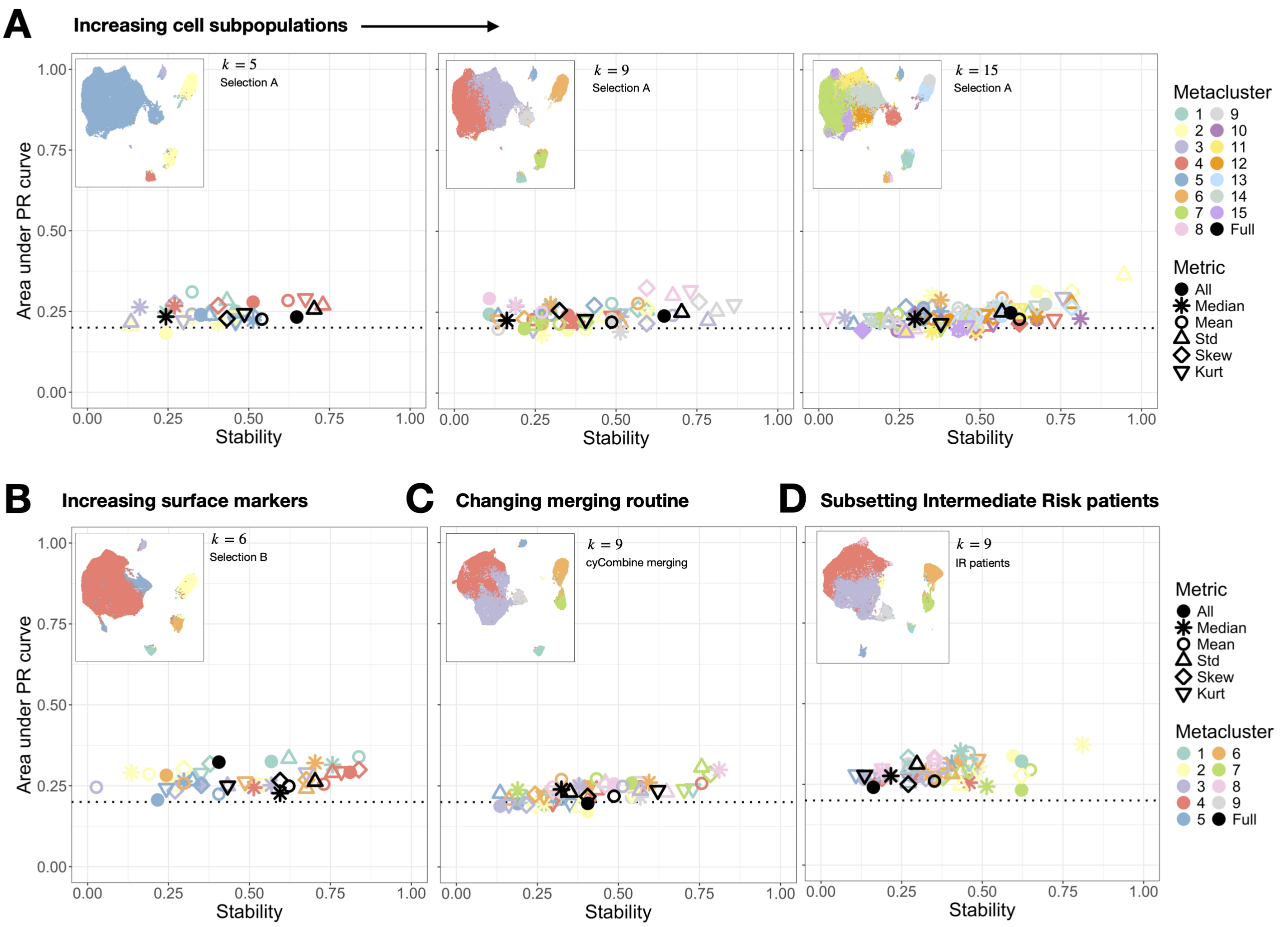


**Figure S20: Reliability assessment of expression-based classification**. Each point represents the performance and stability of the best classifier obtained for each dataset, as discussed in 'Methods' and Figure S6. Color represents the metaclusters (Figure 2) and point shape represents the metric used to summarize the distribution. The interpretation of the AUCPR-Stability plot is as follows: optimal models should reside in the upper-right quadrant, indicating high stability and a large area under the curve. The dashed line represents the average precision of the baseline classifier. **A.** Default results (corresponds to Figure 4C in the main text). **B.** Results for increased number of markers (corresponds to Figure 4D in the main text). **C.** Results for cyCombine merging routine (corresponds to Figure 4E in the main text). **D.** Results for intermediate risk patients (corresponds to Figure 4F in the main text).

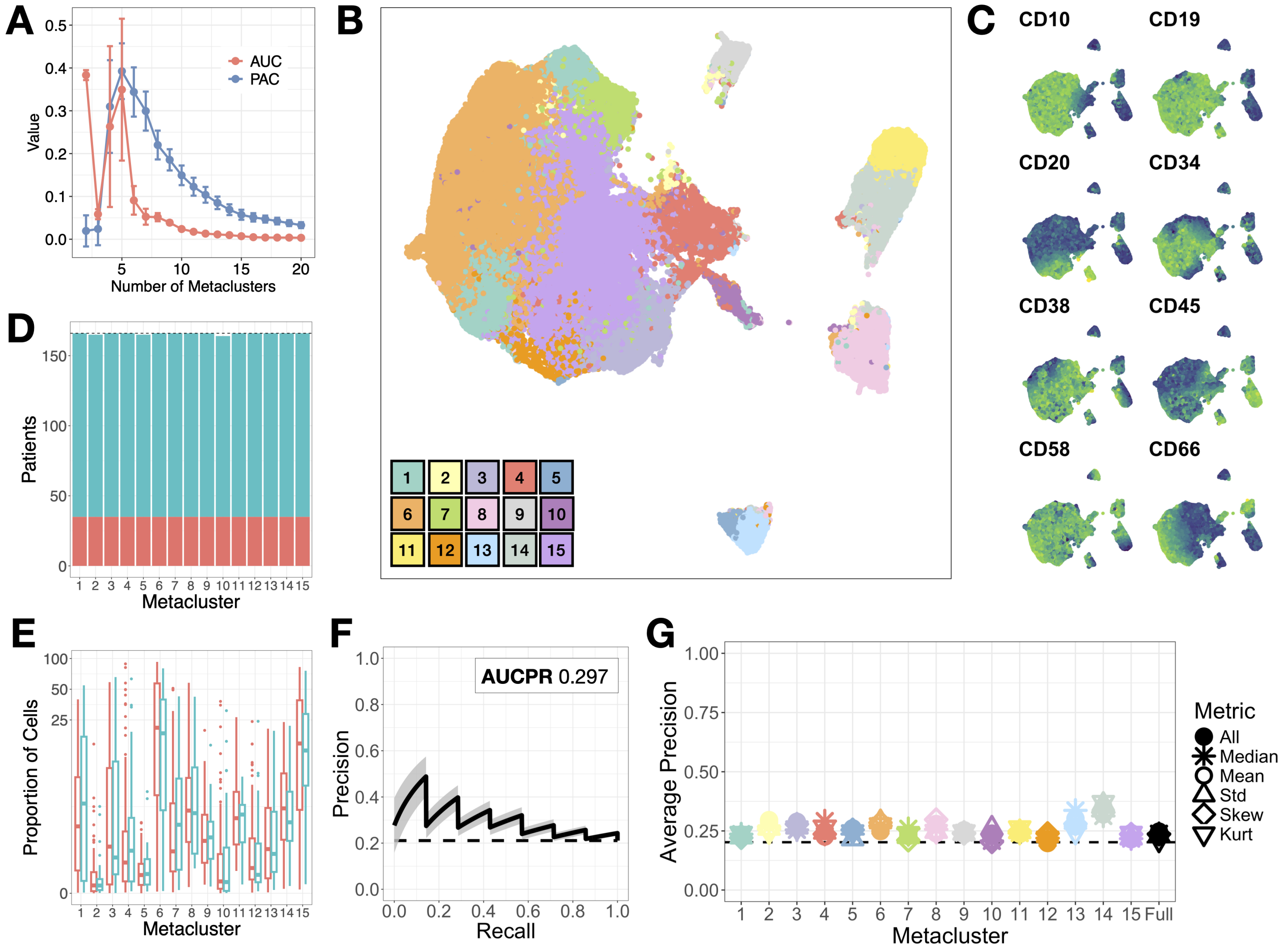


**Figure S21: Results of repeating the analysis with 40000 cells per patient. A.** Values of AUC (Area Under the Curve of the empirical distribution function of a consensus matrix) and PAC (Proportion of Ambiguous Clustering) according to number of metaclusters selected by FlowSOM algorithm. **B.** UMAP visualization of 15 FlowSOM metaclusters. **C.** Relative intensity of marker expression in the UMAP embedding employed for visualization. **D.** Number of patients included in each metacluster. **E.** Comparison of cell percentage per cluster between relapse (R) and non-relapse (NR) patients. Boxplot includes median and IQR. The scale has been transformed with an inverse hyperbolic sine for clarity. **F.** Classification results for abundance in terms of Area Under the Precision Recall Curve using information from all metaclusters. The shaded region represents the standard deviation of 50 repetitions of the classification routine (10 folds + 5 repeats). Horizontal dashed line represents the baseline precision, which equals the proportion of relapse patients in our dataset (Table 1 in main text). **G.** Classification results for expression in terms of AUCPR. Black dashed line represents baseline precision. Color denotes metacluster. Circles represents the average precision obtained when using all the distribution metrics together to train the classifier.

[11] Haidong Yi and Natalie Stanley. Cytoset: Predicting clinical outcomes via set-modeling of cytometry data. In *Proceedings of the 12th ACM Conference on Bioinformatics, Computational Biology, and Health Informatics.* Association for Computing Machinery, 2021.
